# Scalable and comprehensive mosaic variant calling using DRAGEN

**DOI:** 10.64898/2026.02.03.26345450

**Authors:** Sairam Behera, Massimiliano Rossi, Yina Wang, Michal B Izydorczyk, Duke Tran, Clifton L Dalgard, Ester Kalef-Ezra, Kavya Kottapalli, Heer Mehta, Gavin Parnaby, Oona Shigeno Risse-Adams, Sonja W Scholz, Helen Shen, Theodore M Nelson, Arun Visvanath, Xinchang Zheng, Harsha Doddapaneni, Thomas Garcia, Christopher E Mason, Christos Proukakis, James Han, Rami Mehio, Severine Catreux, Fritz J Sedlazeck

**Author notes:** equal contribution (Sairam Behera, Massimiliano Rossi, Yina Wang, Michal B Izydorczyk). equal contribution (James Han, Rami Mehio, Severine Catreux, Fritz J Sedlazeck).

## Abstract

Detecting low variant allele fraction (VAF) mosaic variants without matching controls remains a major challenge in genomics, limited by technical noise, lack of benchmarks, and computational scalability. We present the DRAGEN mosaic caller, a hardware-accelerated approach identifying variants down to ∼1-2% VAF with low false-positive rates and hour-scale runtimes for mosaic SNV/indel detection from bulk sequencing. To support evaluation, we introduce a genome-wide low-VAF benchmark for variants between 1-10% VAF. Application to blood, sperm, and brain tissues revealed patterns, including mosaic hotspots and mutational signatures. The first analysis of HG002 blood showed that many “mosaic” variants defined from HG002 cell lines are likely culture-derived and not in vivo mutations. Importantly, DRAGEN also enables personalized assembly pangenome references to improve alignment and mosaic variant detection in complex regions. Together, this development makes routine low-VAF discovery feasible, opening new opportunities to study mosaic mutations in healthy and disease individuals.

## Introduction

Mosaic variants, the somatic mutations acquired after conception that generate genetically distinct cell lineages, constitute a crucial yet understudied component of human genetic variation with major implications for disease^1,2^. These variants arise post-zygotically and can contribute to a wide spectrum of disorders, including cancer initiation, neuropsychiatric disorders and age-related pathologies, often manifesting years or even decades before clinical symptoms appear^3,4^. Their importance in disease etiology is increasingly recognized, yet detection remains challenging due to their characteristically low allelic fractions (VAF) (typically <1-5%) within affected tissues. Although single-cell sequencing can identify mosaic variants, its limited sensitivity, high cost, and amplification biases restrict its utility for large-scale or tissue-based studies^5,6^. Single-cell data also provides only static snapshots, making it difficult to reconstruct the temporal dynamics of mosaicism^7^. In contrast, high-coverage bulk sequencing (≥80×) can offer a more practical, cost-efficient and scalable alternative, theoretically capable of detecting variants at 1-5% VAF while being compatible with diverse sample types, including formalin-fixed paraffin-embedded (FFPE) tissues. It further was recently shown that bulk sequencing recovers more accurate VAF across tissues than single cell sequencing alone^8,9^.

Current methods for detecting mosaic single-nucleotide variants (SNVs) from bulk sequencing data remain limited, are developed mainly for cancer studies and face substantial scalability challenges. Only a few tools exist (e.g., MosaicHunter^10^, MosaicForecast^11^, and DeepMosaic^12^) that are designed to detect relatively high VAF mosaic variants outside of cancer samples, but these are less optimized for detecting low-fraction (<5% VAF) mosaic SNVs in non-cancerous tissues. Ultra-sensitive somatic callers can reach low VAFs but require ultra-deep sequencing (>500-1,000x) and matched controls, limiting applicability to population studies^13^. Adoption is further constrained by high false-positive rates in low-VAF ranges, necessitating extensive filtering and long runtimes (often days). Most critically, no current method reliably distinguishes true mosaic SNVs from technical artifacts at scale, leaving population-level studies of mosaicism computationally impractical and economically unfeasible with standard bulk sequencing.

A major challenge is the lack of comprehensive genomic benchmarks for low-VAF mosaic variants. There are indeed only a few genomic benchmarks currently available that each summarizes only a few mosaic SNV and indels across their entire genome. A Yonsei University College of Medicine (YUCM) benchmark was recently published, highlighting 354,258 variants across deep sequenced (1,100×) exomes^14^ from an *in-vitro* mixture of six cell lines. This of course is highly limited to exons and excludes complex regions that arguably should harbour the most mosaic variants (i.e., promoters, intergenic regions, and tandem repeats). In contrast, the Genome In A Bottle (GIAB) consortium recently provided a HG002 mosaic benchmark for SNV only^15^. While this is genome-wide, the benchmark is limited in that it only reports 85 SNVs (VAF: 5-30%). However, these are all reported as a minimum of 5% VAF and are only substitutions. Therefore, to enable reliable detection of low-VAF mosaic mutations, we need improved methods and scalable benchmark datasets that include variants below 5% VAF and extend beyond exonic regions. Such resources will be essential for understanding how rare somatic mutations contribute to human evolution and disease.

In this work, we introduce DRAGEN mosaic variant detection as an extension of DRAGEN^16^ unified software framework and systematically evaluate its performance across multiple options, including default and high-sensitivity modes, population and personalized assembly pangenome references, and diverse biological contexts. The new DRAGEN model detects mosaic variants with meaningful sensitivity above ∼2-3% VAF, and partial sensitivity down to ∼1% given high coverage (∼400x or more). We showcase the importance of this innovation by comparing it to other methods and demonstrating the scalability and accuracy of the method across publicly available benchmarks. Nevertheless, given the limitations of many benchmarks (e.g., variants >5% VAF), we further introduce a low VAF-enabled benchmark using a mixture of three GIAB samples that was sequenced and made publicly available. Across all benchmarks, DRAGEN showcases its accuracy and speed to provide variant calls in hours even for datasets at 500x coverage or higher. Thus enabling, for the first time, mosaic mutation ascertainment across all VAF ranges at scale. To further innovate on germline and mosaic detection of SNV and indels, we introduce the concept of a personalized assembly pangenome reference (PAPR). The incorporation of personalized assemblies together with GRCh38 improves variant calling while maintaining GRCh38 coordinates that enable the utilisation of standard annotation resources. Demonstrating this on HG002 cell line and tissues of the individual highlights the improvements compared to a default population pangenome approach, which outperforms the linear reference^16^. We further highlight this across multiple individuals whose tissues we collected and sequenced. These innovations provide unprecedented insights into tissue variability from germline to mosaic scale unlocking new insights into biology and disease research. We highlight new insights across multiple tissues, and further illustrate the capabilities across bulk and single cell WGS from brains of patients with a rare neurodegenerative disease, multiple system atrophy (MSA).

## Results

### Accurate identification of mosaic variants at scale

To enable robust and rapid mosaic variant calling, we extended the framework of DRAGEN^16^ with a new machine learning module to capture low VAF (>1%) mutations from bulk Illumina sequencing. This is now integrated into the existing germline variant calling workflow already capturing SNV, indel, tandem repeats, structural variants, and copy number variants at scale^16^. The challenge and innovation with low VAF mosaic variant calling remains to distinguish noise from actual variants with only limited information available. To accomplish this goal, we i) firstly extracted read level features including statistical descriptions of mapping quality, base quality, strand bias, depth, AF as well as internal Hidden Markov Models (HMMs) scores, SSE (sequence specific error) triggers, and other statistics from DRAGEN variant-calling internal processing; ii) then we trained a gradient-boosted ensemble of decision tree learners using these rich features; iii) applied the model to any evidence loci that’s non-germline for mosaic variant calling (**see methods** for details). **Figure 1A** shows a summary of these steps and highlights the possibility of using a pangenome to aid mosaic variant detection. Our method introduces three major advances for mosaic variant detection: i) Leveraging DRAGEN’s pangenome reference to recover low-VAF variants in challenging genomic regions, ii) A lower evidence threshold to boost sensitivity for subclonal events, and iii) A specialized ML model trained on simulated mosaic data (1-45% VAF) that filters noise while maintaining scalability. Critically, the pipeline requires no matched controls, outputs mosaic calls directly into standard VCFs (tagged for easy interpretation, reported as 0/1 genotypes), and adds minimal computational overhead. In addition to the default mosaic calling, DRAGEN also employs a high-sensitivity mosaic mode (HS) to improve low VAF variant calling. However, we do not recommend using this mode for simple germline calling. By combining DRAGEN’s alignment innovations with a mosaic-trained classifier, we enable accurate, large-scale mosaic detection to become a routine calling procedure.

**Figure 1.**
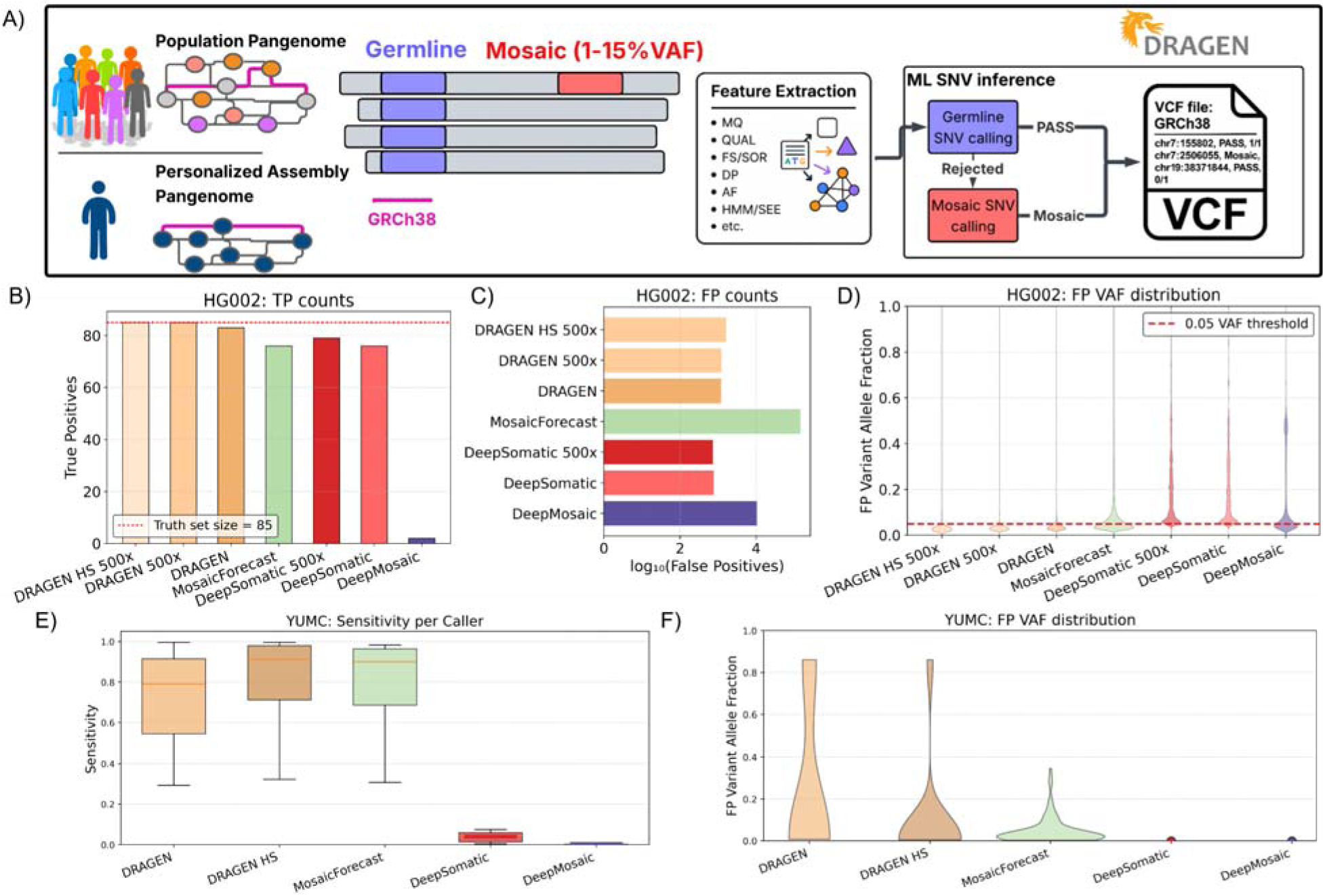
DRAGEN variant calling performance, compared to other popular mosaic variant callers. We utilized 2 standardized benchmarking approaches, a dataset of 85 high-confidence GIAB HG002 SNVs and a mosaic benchmark set from Yonsei University College of Medicine (YUCM). **A)** Workflow diagram of the DRAGEN mosaic variant detection pipeline. **B)** True positive results for HG002 benchmark. DRAGEN detected all 85 variants (red dotted line) at 500x depth of coverage and missed two variants at 80x coverage, due to downsampling reducing the number of informative reads. Additionally, it was run in high-sensitivity (HS) mosaic detection mode. For DeepMosaic, the caller actually identifies 84 out of 85 benchmark variants, however marks 82 as “artifact”, correctly annotating only two **C)** False positive counts for HG002 benchmark. **D)** Variant allele fraction distribution for false-positive HG002 results. MosaicForecast shows the largest number of false-positives, 146,616 with 46.2% above 5% VAF detection threshold, represented by the red line. For DeepMosaic 59.1% (6,254) variants were above the >=5% VAF detection level, while for DRAGEN 15.8% were above detection levels, with only 4.3% in the increased coverage. DeepSomatic shows the smallest absolute FP count (768, 733 at 500x), but all above 5% VAF threshold **E)** Sensitivity results for YUMC benchmark. Per-cell mixture 2,316-9,549 true-positive variants are detectable, with similar patterns across DRAGEN and MosaicForecast, while DeepSomatic detects only up-to 545 TPs and DeepMosaic classifies virtually all calls as artifacts **F)** False positive VAF distribution for YUMC benchmark. Only MosaicForecast has significant numbers of false positives; however, 54% of those variants are under the 5% VAF mosaic detection threshold. DeepSomatic reports 2 FP variants, whereas DRAGEN showed 13 variants across 8 cell mixtures.

We evaluated DRAGEN across three categories of benchmarks: (i) defined GIAB mosaic variants, (ii) the YUCM cell line mixture benchmark, and (iii) a newly created 3-sample HapMap low-VAF mixture covering 1-10% VAF. We further assessed other established variant callers, including MosaicForecast^11^, DeepMosaic^12^, and DeepSomatic^17^. MosaicHunter’s^10^ older design precluded its use, as it could not efficiently process our high-depth sequencing data. It is important to realize that the remaining methods (excluding DeepSomatic) require running Mutect2^18^ first as a variant calling program and then subsequently executing their method. For runtime comparisons, we summed up the time that Mutect2 takes with the individual runtimes from MosaicForecast and DeepMosaic (**see methods**). For DRAGEN, we measured the runtime including the mapping of the reads as the pipeline is highly parallelized and optimized. Similarly, DeepSomatic includes variant calling and annotation in its main pipeline; however, it requires prior alignment to the reference, which we included in the comparison. We report overall F-measure but also list false positives (FP) and false negatives (FN) variants for comparison. Furthermore, for simplicity of comparison we define mosaic variants as being below 15% VAF for the remainder of the paper.

We first benchmarked DRAGEN and other methods on HG002 GIAB that recently postulated 85 mosaic mutations ranging from 5.5% to 27.2% in VAF^15^. On a ∼500x Illumina whole genome run, DRAGEN identified all 85 variants within 12 hours 2 minutes of runtime (from raw reads to VCF file). DeepSomatic was also able to analyse the 500x coverage sample, recovering 79 variants and reporting only 733 false-positives in 3 hours 4 minutes; however, it is important to emphasize that running DeepSomatic required alignment of the reads (using BWA-MEM2) to the reference genome prior to variant calling and classification, which took 89 hours 4 minutes, bringing total processing time to 92 hours 8 minutes. In contrast, other methods were unable to finish the analysis within a week of runtime even without the mapping step included. Therefore, we subsampled the data to 80x coverage. With this coverage, DRAGEN was able to report 83 out of the 85 variants (**Figure 1B**) where the missed variants showed biases introduced during downsampling (**Supplementary Figure S1**). Upon closer investigation DRAGEN rejected one true positive (TP) as it showed a heavy strand bias in the subsampled data. Moreover, DRAGEN showed low additional variant calls, reporting only 1,212 variants absent from the benchmark set, while it found 1,232 at 500x (1,627 variants in HS detection mode, **Figure 1C**). In comparison, MosaicForecast was able to correctly detect and annotate 76 TP mosaic variants present as part of the benchmark. However, in addition, it reported 146,616 variants that were absent from the germline HG002 calls. DeepSomatic similarly found 76 TP variants but reported the lowest false-positive (FP) rate of only 768 variants. DeepMosaic reported 84 of the variants present in the benchmark set, however, labeling 82 TP variants as artifacts. In addition to these, DeepMosaic reported 399,883 false-positive variants, while the majority (389,301) were labeled as artifacts (**Figure 1C**).

Given the GIAB benchmark only reported variants down to 5% VAF variants, we next assessed if the potential FP calls are below that VAF threshold. Analysis of the FP variants reported by DRAGEN shows that only 53 FPs are above the 5% VAF threshold at 500x coverage (4.3% of total FPs), with 103 (6.3%) FPs with high-sensitivity mosaic detection mode enabled, whereas for the 80x sample, 191 FP are above the 5% VAF threshold. Other callers show a higher proportion of FP above 5% VAF, with DeepMosaic at 59.1% (6,254 variants) and MosaicForecast at 46.2% (67,729 variants). Interestingly, all FP reported by DeepSomatic are above 5% VAF (**Figure 1D**). Thus, highlighting that DRAGEN has the highest TP and lowest FP for all variants called above 5% VAF, which was the reported threshold for GIAB^15^. A key advantage of DRAGEN is its runtime; it completed all the processing in 12 hours (500x coverage), while MosaicForecast and DeepMosaic are 7.5 and 7.7 times slower, as they depend on running BWA and Mutect2 before performing variant classification. However, DeepSomatic was able to perform variant calling on the downsampled HG002 in 2.25 hours (50 hours including mapping with BWA) by running each chromosome concurrently on separate compute nodes; processing the whole genome on a single compute node would take 24.6 hours (**Supplementary Table S1**).

To further assess DRAGEN, we next utilized the benchmark proposed by researchers at the Yonsei University College of Medicine (YUCM)^14^, which was generated based on MosaicForecast ^11^ and DeepMosaic ^12^. The benchmark is based on 1,100x whole-exome sequencing of eight specific cell mixtures, designed to simulate a spectrum of mosaic variants. Here, DRAGEN remains by a distance the fastest method, simultaneously being able to identify the majority of the variants among the examined cells, reporting up to 9,549 variants per cell line mixture and missing only 36 variants in the same sample (**Supplementary Table S2**). Utilization of high-sensitivity mosaic detection mode improves the results even further as it enables detection of 836 additional TP YUMC benchmark variants on average across samples. In contrast, other callers often missed twice as many, reporting up to 9,501 variants per cell line, with MosaicForecast missing 131 variants and DeepMosaic missing 84. However, this varied sample to sample, as average recall for DRAGEN was 0.713, with standard deviation (SD) of 0.273. High-sensitivity mode increased the recall to 0.812 and reduced SD to 0.237; for DeepMosaic the average recall/SD was 0.691/0.275 while for MosaicForecast it was 0.794/0.239 (**Figure 1E**). Misclassification was again a large issue for DeepMosaic; in the YUCM dataset it flagged virtually all of its findings as artifacts. DRAGEN and DeepMosaic showed minimal false-positives (0-5/0-4 variants per cell), while MosaicForecast exhibited slightly higher values of 5-14 FP variants per cell mixture. Nevertheless, the precision for all three callers was above 99.4%. In comparison, DeepSomatic was the least sensitive method, recovering on average 310 TP (recall/SD: 0.040/0.028) but thus naturally also had the lowest number of FP (**Figure 1F**).

Next, to assess the accuracy of mosaic variant detection down to 0.75% VAF, we prepared an admixed sample by combining genomic DNA from three well-characterized NIST GIAB control samples (HG001, HG002, and HG005), for which curated SNV benchmark (GIAB v4.2.1) sets are available^19^ (**see Supplementary Section 1**). The DNA was combined in a ratio of 60:2:1, corresponding to 95.24% HG005, 3.17% HG001, and 1.59% HG002 (**Figure 2A**). This mixture mimics a combination of germline and mosaic variants, as the contributions from HG002 and HG001 result in lower VAFs, while HG005 contributes to higher VAF germline variants. The expected VAFs for homozygous variants were 3.17% for HG001 and 1.59% for HG002, whereas the expected VAFs for heterozygous variants were 1.59% and 0.79% for HG001 and HG002, respectively (**Figure 2B**). Leveraging the GIAB benchmark sets for these three individual samples, we constructed an admixed benchmark set (see **Methods**). In this set, variants from HG005 were considered germline variants, while variants found only in HG001, HG002, or shared between them were designated as mosaic variants (**Figure 2C**). The distribution of benchmark mosaic variants based on the HG001 and HG002 genotype combination is shown in the **Supplementary Figure S2**. Altogether, this benchmark includes 3,512,786 germline and 2,208,603 mosaic variants. The benchmark covers 80.12% of the human genome (GRCh38). The benchmark is available at https://github.com/michalizydo/dragenMosaic.

**Figure 2:**
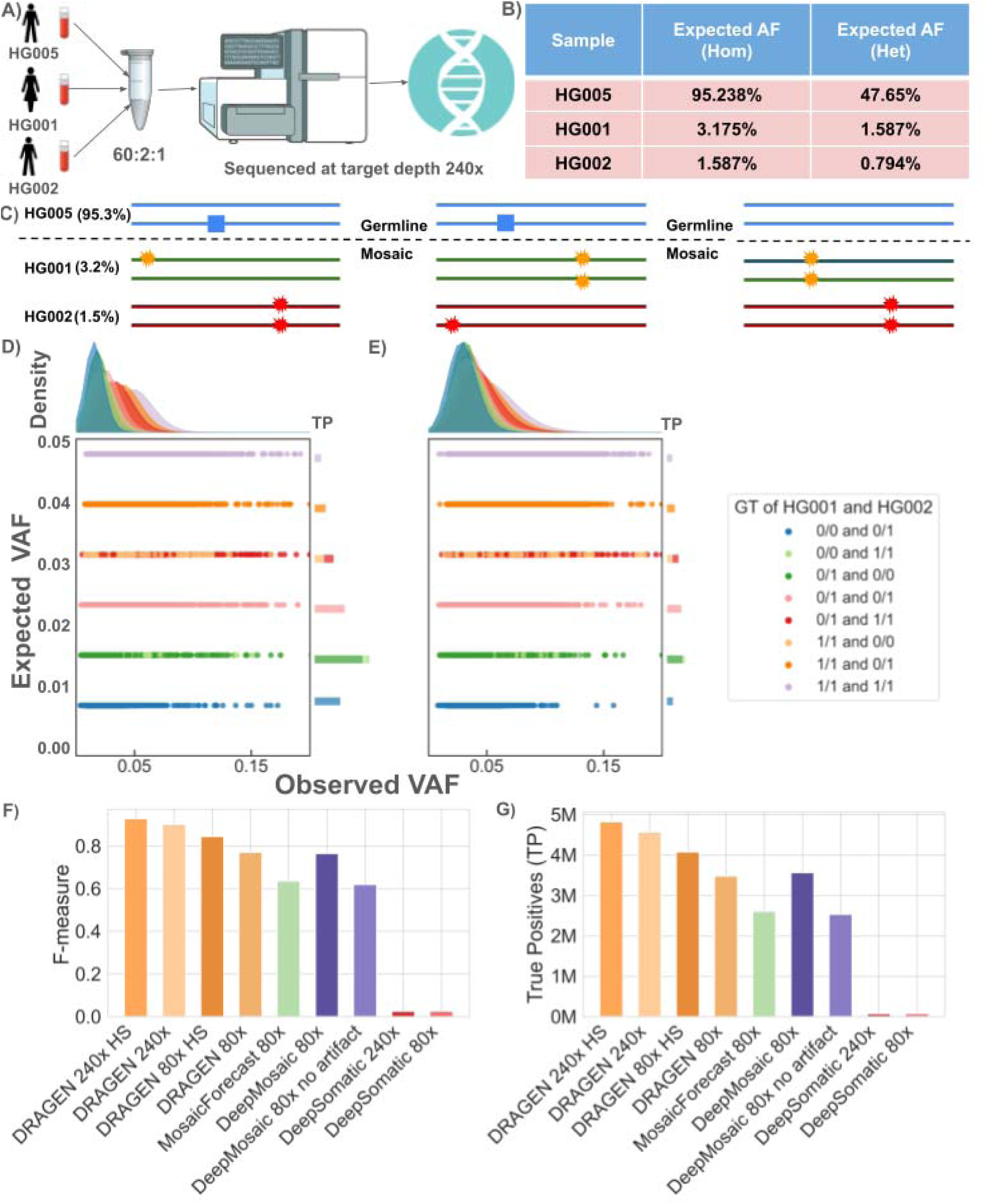
HapMap mixture: **A)** Three GIAB samples HG005, HG001 and HG002 were mixed in-vitro with 60:2:1 ratio and sequenced at 240x coverage **B)** The expected variant allele fraction (VAF) of the variants in this mixture **C)** Mosaic variants based on their presence in HG001 and HG002 **D)** Expected VAF vs Observed VAF for true positives of 240x dataset; the distinct horizontal clusters corresponds to the mosaic variants with different genotype (of HG001 and HG002) combinations. The density plot above shows the overall distribution of observed VAFs and the right-side bars shows the true positive mosaic variant numbers corresponding to the genotype combinations of HG001 and HG002 and **E)** downsampled 80x dataset of HapMap sample **F)** Comparison of TP of different methods **G)** Comparison of F-measure. DRAGEN was run with both default and high-sensitivity mosaic detection mode; also on 240x dataset.

To assess DRAGEN’s accuracy for detecting low-VAF mosaic variants, we sequenced an *in vitro* mixture of three samples to 240x Illumina whole-genome coverage. At this depth, DRAGEN identified nearly 4.72 million variants in total, of which ∼795,000 had VAF ≤ 15%. Among all these low VAF (<=15%) variants, approximately 90% are below 5% VAF with an average VAF of 0.034. Benchmarking all germline and mosaic variants against our benchmark set demonstrated strong performance, yielding an F-measure of 90.14% (sensitivity: 82.15%, precision: 98.84%). We also ran DRAGEN with its high-sensitivity mosaic detection mode that recovered more TP, improving the overall sensitivity to 86.78% and F-measure to 92.78% albeit slightly decreasing precision (98.68%) (see **Supplementary Table S3**). Next, we evaluated the sensitivity of mosaic (VAF <= 15%) variants based on their expected VAF. The highest recall (84.4%) was achieved for an expected VAF of approximately 4% and results around 3-5% showed comparable sensitivity (74-78%). Recall decreased with a decrease in expected VAF, falling to ∼60% near 2% VAF and below 35% for variants under 2% VAF, with the lowest detection (∼6%) observed at 0.8% VAF (see **Supplementary Table S4 and Supplementary Figure S3**). This is likely due to insufficient ALT coverage, as the lowest VAF is only represented by two reads on average. Overall, observed VAFs matched the theoretical expectations, with most true alleles deviating by less than 0.02 from their expected fractions. For all genotype combinations corresponding to HG001 and HG002, the observed VAFs showed prominent peaks below 0.05 that confirms the moderate but significant positive correlation (pearson r=∼ 0.70, p-value = 0) between observed and expected VAFs (shown in **Figure 2D**). Together, these findings indicate that DRAGEN reliably detects the majority (>70%) of true mosaic variants above 3% VAF, but sensitivity diminishes rapidly for lower VAF alleles like due to coverage limitations. Out of other tested mosaic callers, only DeepSomatic was able to produce a call set for this mixture at 240x coverage, while the other callers took over a week of runtime. However, while maintaining the lowest FP number of 3,873 variants, DeepSomatic detected only 65,143 variants from the benchmark, with an overall recall of only 1.2%. This is likely due to the majority of the benchmark SNVs being below the 5% VAF range.

We subsampled the HapMap mixed data to 80x coverage to enable a direct comparison between DRAGEN, MosaicForest, DeepMosaic, and DeepSomatic. As expected, the low VAF variants in the benchmark are not easily detected at 80x, and thus, we observed an overall reduced sensitivity (**Supplementary Table S5**). The observed VAFs showed a modest positive correlation (pearson r = 0.47, p-value = 0) with the expected VAFs for all different genotype combinations (**Figure 2E**). The density plots also showed that the peaks for observed VAFs closely aligned with the expected values, indicating only minimal deviation among the true positive mosaic variants. This effect was evident in DRAGEN, where TP variants dropped from 4.33 million at 240x to 3.71 million at 80x (**Figure 2F**), and thus the F-measure reduced from 85.68% to 78.25%. Utilizing DRAGEN’s high-sensitivity mosaic detection mode improved detection at both coverage rates, reporting 4.95 million TP variants at 240x and 4.19 million TP at 80x, with an F-measure of 92.17% and 84.02% respectively. In comparison, MosaicForecast at 80x coverage identified 2.65 million TP, with F-measure of 62.68% (∼21% less than DRAGEN). DeepMosaic showed intermediate performance, calling 3.63 million TP but generating a high number of FP (265k), leading to an F-measure of 75.44%. However, filtering out variants flagged as artifacts reduced TP to 2.56M, FP to 102K, and F-measure to 61.18%. DeepSomatic detected only 71K TP and 7.8K FP at 80x, showing similar performance regardless of the tested sample coverage. Overall, these results underscore that DRAGEN remains the most accurate method across conditions (**Figure 2G**). Although DeepMosaic and MosaicForecast achieved relatively high raw detection counts, DeepMosaic reported only 3% less total counts than DRAGEN at the same coverage and their results were heavily affected by “artifact” misclassification and FP.

Thus, across all three benchmarks, we could demonstrate that DRAGEN mosaic variant calling performs accurately even below 5% VAF, while providing results within hours of runtime on high coverage data sets without matching controls. This innovates mosaic variant calling from bulk sequencing data at scale and accuracy.

### Mosaic mutation detection in brain tissue for MSA patients

We next applied DRAGEN to investigate mosaic variation in MSA brain tissue (**Supplementary Figure S4**). MSA is a neurodegenerative synucleinopathy with unclear aetiology and low heritability; mosaic mutations confined to small cell populations may promote prion-like protein spread and contribute to disease, with mosaic CNVs of the alpha-synuclein gene already shown to be related to pathology^20^. We compared DRAGEN calls across two MSA patients’ brains that have available 85x Illumina whole genome bulk samples and multiple single-cell libraries (16 single cells for MSA1 and 15 single cells for MSA2 brain, average coverages 0.8x and 1.1x, respectively)^21^. These were generated using droplet Multiple Displacement Amplification (dMDA), an isothermal method which has a low per-base error rate^21^. Given all the advantages and disadvantages of single cell sequencing mentioned above, we were curious about how bulk sequencing with the DRAGEN analysis compares to the single-cell sequencing of the same region across these two brains.

We thus compared bulk mosaic variants VAF with their presence across single cells (**Figure 3A**). DRAGEN identified an average of ∼38,000 mosaic and ∼5,000,000 germline variants per bulk sample. Using bulk-identified mosaic variants as targets, we interrogated up to 13,890 loci per brain in single cells, genotyping 849-5,887 variants per cell, with 130-1,093 ALT alleles detected, which is consistent with sparse single-cell representation of mosaic sites. Across the samples of both brains, the fraction of single cells carrying an ALT allele increased steadily with bulk VAF (Spearman ρ = 0.879, p < 10□□, **Supplementary Figure S5**), confirming the reliability of DRAGEN mosaic calls. Notably, most variants clustered near the 0.15 VAF cutoff used for bulk mosaic detection, with ∼20 % of single cells supporting these low-VAF variants. A smaller subset showed near-ubiquitous presence across cells, suggesting either clonal enrichment or misclassification of low-fraction germline variants. Overall, 76.3% of bulk mosaic variants were genotyped in at least one single cell, providing strong orthogonal support for DRAGEN mosaic calls while revealing biological heterogeneity among low-VAF sites.

**Figure 3:**
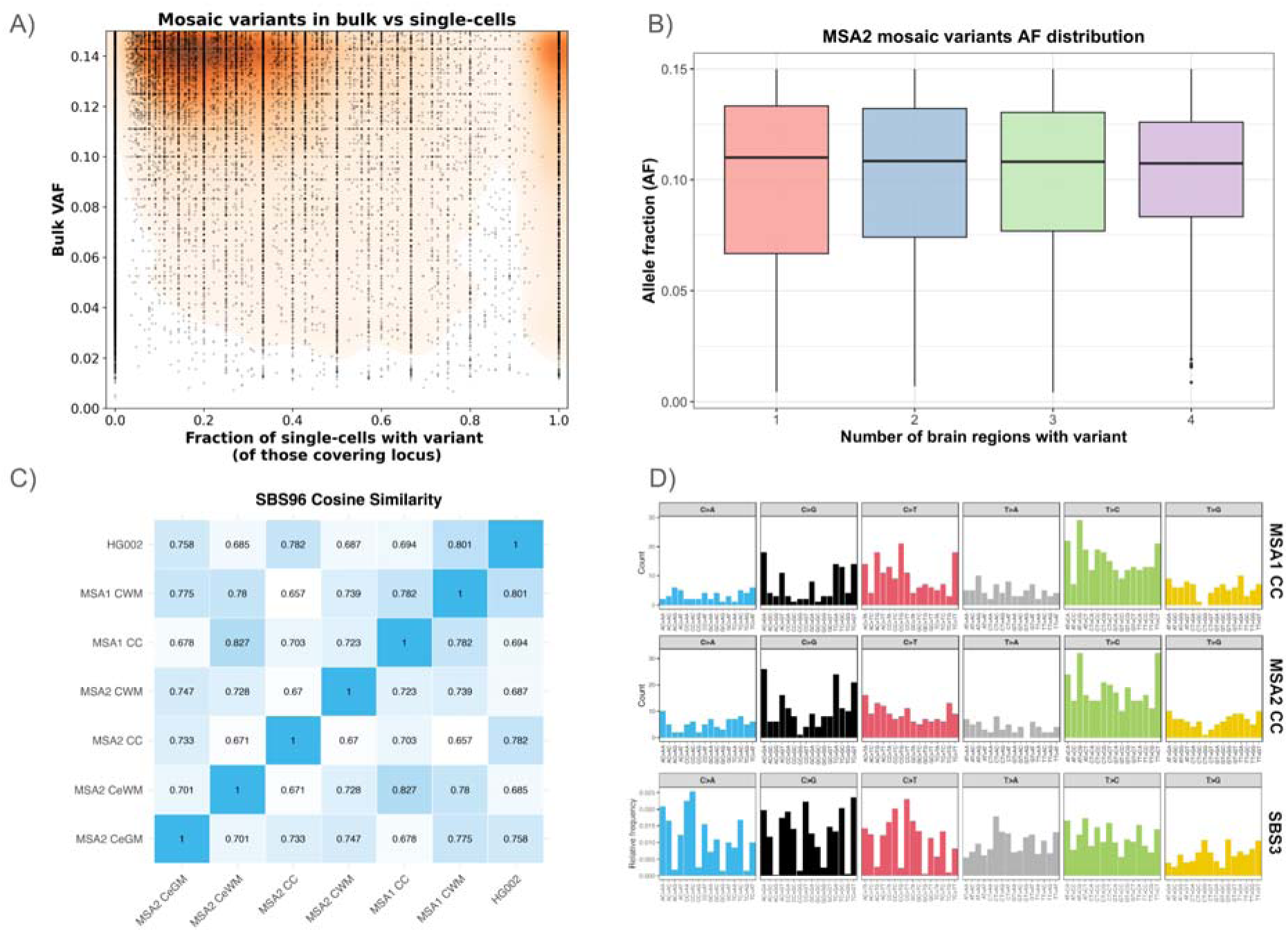
Mosaic variants across the brains. **A)** Density map of mosaic variants found in bulk and their presence in corresponding single cells. Data was aggregated between two studied MSA brains. The concentration is highest towards the 0.15 bulk VAF cutoff for mosaic; however variants with VAF as low as 0.01 in bulk are detected. The majority of recovered variants are distributed in three peaks and represented by 4 single cells carrying the ALT allele, with 12, 8 and 4 cells covering the locus, respectively. **B)** Overlap of the mosaic variants in the MSA2 brain. Roughly half of all mosaic variants detected in cingulate regions are also present in cerebellar regions, whereas only about 40% of cerebellar variants overlap with the cingulate. Values in brackets represent the average VAF of mosaic variants in each set. Per-region variant overlap for both brains is included in **Supplementary Figure S6 and Supplementary Figure S7**. **C)** Cosine similarity of SBS96 variant mutational patterns across the bulk brains, with only <5% VAF mosaic variants included. **D)** SBS96 mutational spectra of cingulate cortex regions of the studied MSA brains, showing significant cosine similarity to SBS3 COSMIC pattern, which results from defective homologous recombination-based DNA damage repair, and was not reported in aging neurons and oligodendrocytes^32^.

In the first MSA brain, which included analyses of the cingulate gyrus cortex and white matter, roughly 30% of mosaic variants (5,083) were shared between these two regions (**Supplementary Figure S6**). Mosaic variants detected in a single brain region and those shared across both regions exhibited very similar allele-fraction distributions (median VAF ∼0.1 within the 0-0.15 range). Variants present in both regions did not show a systematic shift toward higher VAF compared to region-restricted variants, indicating that broader spatial spread of mosaic variants is not strongly associated with higher clonal fraction in this brain (**Supplementary Figure S7**). The other brain, for which the cerebellar gray and white matter were also sequenced, shows pronounced regional differences in the distribution of mosaic variants. Cingulate cortex and cingulate white matter share a large fraction of their mosaic variants, with relatively few mutations confined to either compartment, confirming a broadly homogeneous mutational landscape across cortical and subcortical cingulate tissue.

When summing all intersections that contain both cingulate regions (i.e., variants present in both cingulate cortex and cingulate white matter, irrespective of whether they were also detected elsewhere), we observed a total of 4,975 shared variants. The corresponding total for variants shared between the two cerebellar regions (gray and white matter) was 4,911. Each region contained 29,600 and 28,531 mosaic variants in total, respectively. Thus, the overall degree of sharing between gray and white matter was comparable in the cingulate and cerebellum, representing roughly one-sixth of each region’s mosaic variant complement. Both pairs also harbored extensive sets of region-specific variants that did not overlap with those in other tissues, consistent with the strong anatomical compartmentalization evident across the overlaps (**Supplementary Figure S8**). Mosaic variants present in one, two, three, or all four regions showed very similar VAF distributions, with median values consistently around ∼0.1 within the 0-0.15 VAF range. There was no clear trend toward higher VAF in variants shared across more regions, indicating that in this brain, the clonal fraction of a mosaic event is largely decoupled from how broadly it is anatomically distributed (**Figure 3B**).

To gain insight into the biology of the mosaic variants found in the brains, we performed mutational signature analysis of SNVs/InDels reported under 15% VAF in the bulk brains, taking advantage of the fact that we had deep WGS from several regions. The initial pattern was very similar across the tissue within the brains, as well as between the two brain samples and HG002 control (**Supplementary Figure S9**). This attested to DRAGEN consistency across different samples. To gain deeper insights, we next examined variants with VAF ≤ 5%, focusing on the most likely later onset mutations. Confirming this, we observed differences between HG002 and the brain samples highlighted in **Figure 3C**. Interestingly, a number of mutational patterns reported in the brains show similarity to standard panels of COSMIC spectra resulting from defective homologous recombination-based DNA damage repair (**Figure 3D**), as well as DNA mismatch repair and microsatellite instability^22,23^. To further investigate the functional aspect of the mutations, we analyzed the mutational burden on the genome and if it targets any functional regions involved in neurodegenerative disorders. Using a 10 Mbp sliding window (see methods), we identified an average of 752 mutational hotspots per brain, with peak densities exceeding 1,900 variants per window. In contrast, the overall genome-wide average was only 49 variants per window. A gene ontology analysis of the hotspot gene list highlighted 27 affected genes, 6 of which are associated with neurodegeneration (see methods, **Supplementary Table S6**). Notably, these genes are reported to impact neuronal and ion channel functions, including *CACNB2, CHRNA7,* and *CLCC1*, which have been implicated in neuropsychiatric and neurodegenerative conditions such as autism, epilepsy, and Alzheimer’s-like syndromes ^24–26^. Additional candidates such as *ACOX1* and *NOTCH1* are linked to oxidative stress responses and neuronal ageing, respectively ^27–30^. While these associations remain exploratory, their recurrence across mosaic-enriched regions highlights intriguing connections between neuronal homeostasis, oxidative damage, and mosaic genome variation in the brain^31,22^.

Together, the MSA analyses show that low-VAF mosaicism is widespread across brain regions. Although all regions display similar mutational signatures consistent with DNA mismatch repair defects, each region also harbors a substantial number of unique variants that are not observed in other analysed brain tissue samples. In addition, several recurrent variants cluster in genomic hotspots, including genes implicated in neurodegenerative diseases. These findings demonstrate that bulk mosaic detection using DRAGEN can recapitulate patterns observed in single-cell data while highlighting candidate genomic hotspots potentially relevant to neurodegeneration.

### Utilizing a personalized assembly pangenome references for variant detection

Low-VAF variants are disproportionately enriched in repetitive and structurally complex regions^11,33^, where linear reference genomes perform poorly^16,34^. Although pangenome references reduce misalignment, they primarily capture population-level diversity rather than individual haplotypes^35^. The personalized assembly pangenome reference (PAPR) incorporates a phased, sample-specific diploid assembly into a pangenome reference while retaining GRCh38 as the coordinate backbone. PAPR can be used in conjunction with DRAGEN as this design enables variant calling to be informed by an individual’s resolved haplotypes, while allowing all variants to be projected back onto the standard reference genome, thereby overcoming key limitations of current approaches that rely on mapping exclusively to phased assemblies and enabling continued use of established annotations and downstream resources.

To evaluate the impact of this approach, we compared DRAGEN’s default pangenome reference (256 haplotypes)^16^ to a PAPR constructed by integrating a sample-specific phased diploid assembly alongside GRCh38 (**Figure 1A**). Thus assessing if a PAPR approach can improve upon an already comprehensive human population pangenome. In theory, a PAPR could improve the variant calling of germline and mosaic SNV and Indel as all germline variants are already present, including more complex variations (e.g. HLA)^36^. As a validation step, we first assessed PAPR performance using established germline variant benchmarks to confirm correctness and baseline behavior of the PAPR configuration. Having established this foundation, we then evaluated the impact of PAPR on mosaic variant detection.

To assess the potential of the PAPR, we utilized the phased genome assembly of HG002 T2T Q100 v1.1 ^37^ and GRCh38 to build our own PAPR representing HG002 (**see Methods**). Next, we aligned Illumina data from the HG002 GIAB cell line (∼500x coverage) to the default and HG002 PAPR to assess the performance of DRAGEN on germline and mosaic SNVs and Indels. The variant calling process, beginning from the alignment step, required approximately 21 hours to complete. Genome-wide benchmarking with the GIAB Q100 v1.1 benchmark set showed that the PAPR improved germline variant calling relative to the default pangenome (F-measure = 99.95% vs 99.85%), yielding 4,520 (8.68%) less false negatives (FNs) but 684 (3.01%) more false positives (FPs) and reducing the overall errors by 5.18%. Consistent results were observed for the GIAB v4.2.1 benchmark (F-measure = 99.86% vs 99.82%), where the PAPR identified 3,412 (31.01%) fewer FNs but only two additional FPs (**see Figure 4A and Supplementary Table S7 and Supplementary Figure S10**) with overall 22.55% less FP+FN. Among the FP variants in the GIAB v4.2.1 benchmark detected by both references, 2,644 were shared, while 494 were unique to PAPR and 205 to the default pangenome. Notably, a small fraction of these FPs (18 in the PAPR and 20 in the default) were labeled as false only due to genotype mismatches. In these cases, the same variant position and alleles were present in both callsets, but the zygosity differed. Furthermore, 301 (9.59% of total FPs) of the PAPR FPs and 95 (3.33% of total FPs) of the default FPs were actually classified as TP in the GIAB T2TQ100 benchmark. Thus, indicating that several of these FP calls reflect differences in benchmark labeling rather than genuine errors. Thus, likely initially missed in the GIAB v4.2.1 benchmark. The PAPR also improved performance in the challenging medically relevant gene regions (CMRG v1.0^38^), achieving an F-measure of 98.98% compared to 98.79% for the default pangenome (**Figure 4B**). This corresponded to 81 (19.38%) less false negatives but only two (2.15%) more false positives. Overall, this highlights the benefit of a PAPR for germline variant calling.

**Figure 4:**
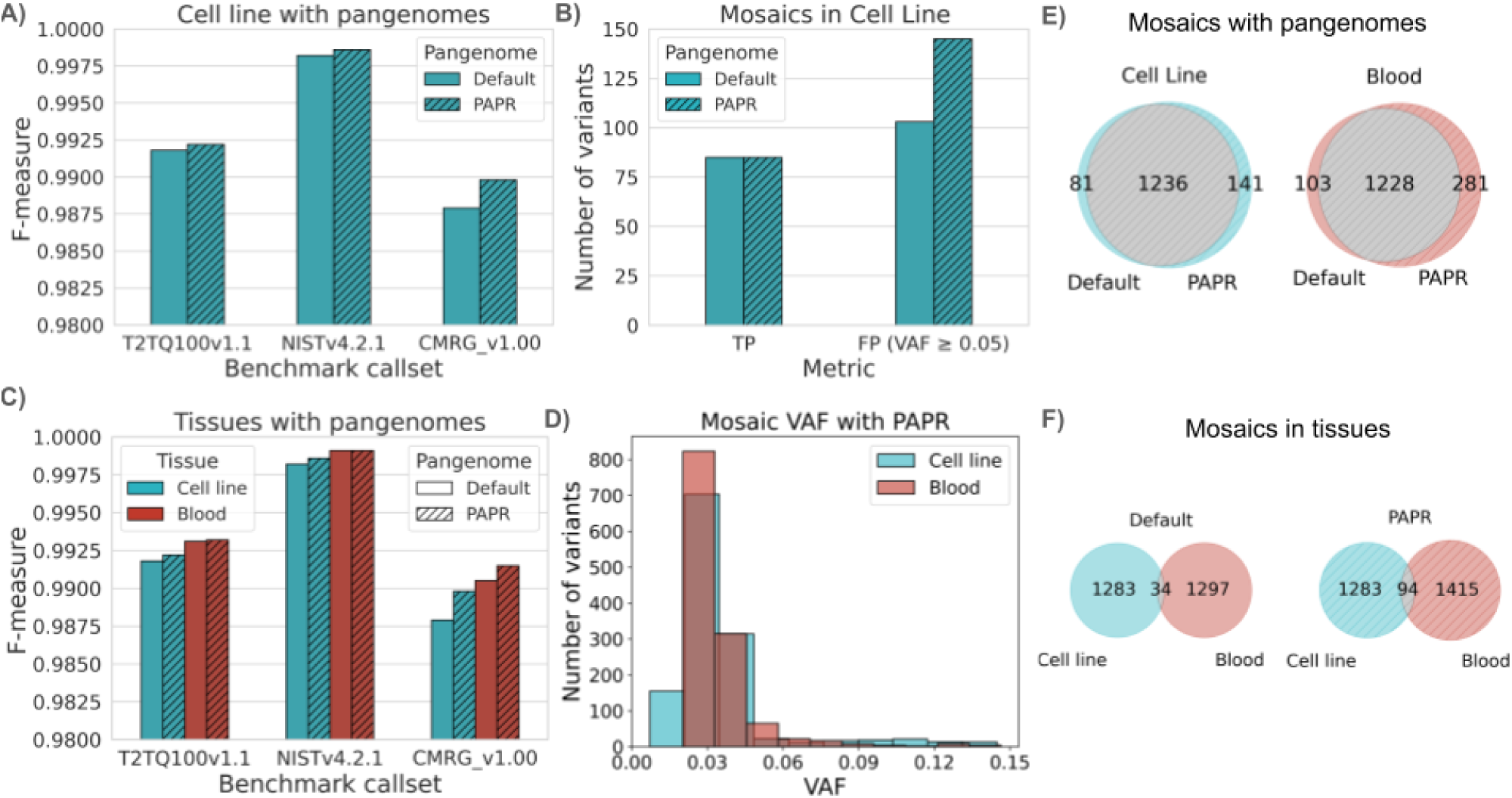
Benchmarking and mosaic analysis with personalized assembly pangenome reference: **A)** HG002 cell line genome-wide benchmarking using T2T Q100 v1.1-v0.019, GIAB v4.2.1 and CMRGv1.00 benchmark sets for cell line with both default and PAPR pangenome **C)** True positive (TP) and False positives (FP) for GIAB mosaic v1.1 benchmark with two different pangenomes. The FPs are filtered with VAF >= 0.05 as GIAB benchmark is restricted to VAF=0.05 **C)** Benchmarking of both cell line and blood with all different benchmark sets and the two different pangenomes **D)** Variant allele fraction (VAF) distribution of mosaics in cell line (∼500x) vs blood (∼100x) with the PAPR built using HG002 T2T Q100 v1.1 assemblies. The bin size 0.015 was used for this plot. **E)** The comparison of mosaic variant counts among default and PAPR for blood and cell line separately. **F)** The comparison of SNV counts among cell line and blood for different pangenomes.

Next, we investigated the mosaic variant calling impact on the two pangenome references (**see Methods**). First, we compared the variant calling performance to the postulated 85 mosaic variants from the GIAB v1.1 mosaic benchmark set ^15^. Here, we found that both the PAPR and default pangenomes were able to detect those variants (**Figure 4B**). In addition to these 85, we identified 53 putative FP mosaic variants (5%<=VAF<=15%) for the default pangenome and 116 for the PAPR in the high-confidence regions of the GIAB mosaic v1.1 truthset, with 35 common to both the pangenomes (see **Supplementary Table S7**). The GIAB v1.1 mosaic benchmark reports variants only down to a VAF of 5%. Consequently, a large fraction of the additional variants detected by both approaches (1,179 using the default reference and 1,176 using PAPR) fall below this VAF threshold and are therefore not represented in the benchmark. In addition, 35 mosaic variants with VAF > 5% were identified by both pangenome references but are absent from the NIST GIAB callset and are thus classified as FP; several of these sites were further examined using IGV and showed visual evidence consistent with mosaic variation (**Supplementary Figure S11**).

To further explore the origin of previously reported mosaic variants and distinguish true mosaic events from cell line artifacts, we collected and sequenced native blood tissue from HG002, marking the first time that in vivo tissue from this individual has been analyzed, whereas prior benchmarks have relied exclusively on immortalized cell line DNA. The blood genome was sequenced to 100x Illumina coverage and analyzed using both the default and PAPR reference.

To our surprise, when benchmarked against the GIAB v4.2.1 dataset for the default pangenome, the blood sample detected 5,918 more TP variants than the cell line genome (∼500x coverage), resulting in slightly higher overall performance (F-measure 99.91% vs. 99.82%). Similarly, benchmarking with the CMRG identified 156 additional TP variants in these difficult regions for the blood tissue (see **Figure 4C and Supplementary Table S7**). These differences might originate from a more recent and more harmonized sequencing run of the blood sample at our center compared to 500x combined data sets from the cell line sequencing.

We next analyzed mosaic variants (VAF ≤ 15%) directly in the blood sample and also compared them with the cell line. The VAF distribution among cell line and blood was different (**Figure 4D**) with more mosaic variants in cell line being below 5%, likely due to higher coverage of the cell line sample. Using the default pangenome for the blood sample, we identified 1,331 SNVs and indels (mean VAF = 0.0505), including 1,171 variants with VAF <5%. The PAPR detected 1,509 mosaic variants (mean VAF = 0.1018), with 1,171 below 5% VAF. Of these, 1,228 variants (mean VAF = 0.0417; 1,116 below 5% VAF) were shared between both references, underscoring strong concordance while revealing subtle differences in low-VAF detection. On the other hand, fewer mosaic variants were identified in the cell line with both the pangenomes (1,232 with default pangenome, 1,291 with PAPR). We also observed a similar trend in both cell line and blood for shared mosaic variants among these two pangenomes (**Figure 4E and Supplementary Table S8**). The higher number of mosaic variants in the cell line compared to blood was due to the high coverage datasets. Comparison of mosaic calls between blood and the corresponding cell line showed limited overlap, with only 34 shared variants for the default and 95 for the PAPR (**Figure 4F**).

We next compared these variants over the GIAB mosaic benchmark variants to assess their biological relevance. None of the 85 mosaic benchmark variants were detected in blood at 100x coverage, and force genotyping revealed no supporting reads for 84 sites and only a single read (VAF = 0.008) for one locus. These results suggest that 84/85 canonical GIAB mosaic variants are likely cell-line-specific variants rather than in vivo mosaic mutations. When comparing the HG002 blood sample to the cell line further, we indeed only found limited overlap. This is consistent with GIAB’s finding that these mosaic variants differ between cell line passages, so that sequencing the NIST Reference Material DNA batch is important for using this benchmark for mosaic variants. These differences highlight biologically meaningful divergence between the cell line and primary tissue genomes, suggesting that immortalization or extended culture may introduce variants absent from the donor’s mosaic tissues.

Together, these results demonstrate that PAPR consistently improves recall and precision on germline variant calling and also showed potential benefits in mosaic variant calling, especially in medically relevant and structurally complex regions.

### Mosaic mutations across blood & sperm utilizing a personalized assembly pangenome reference

Given the improvements of the personalized assembly pangenome reference (PAPR) on HG002, we next evaluate its feasibility in the absence of a perfect T2T assembly (such as Q100). Furthermore, we assessed PAPR outside of the cell line (such as HG002) on tissues from individuals that donated blood and sperm. The accessibility of these tissues allows systematic measurement of mosaic variants within individuals across distinct cellular lineages. For this, we sequenced blood and sperm samples from three individuals at approximately 100x coverage each. For the blood samples, haplotype assemblies were generated using PacBio HiFi datasets, and subsequently, PAPRs were constructed for each individual (**see Methods**). We then performed variant calling for each sample using the corresponding PAPR reference built from the blood assembly.

For all germline and mosaic variants within the GIAB regions across autosomes, we first compared three samples separately for blood and sperm to examine how variants are shared among individuals. In blood, we identified 2,024,902 variants common to all three samples (∼50% of the total per sample), an average of 861,092 variants unique to each sample, and about 536,659 variants shared by any two samples (see **Supplementary Table S9**). A nearly identical pattern was observed in sperm, with 2,024,975 variants common across all samples, an average of 861,121 unique variants per sample, and 536,543 variants shared by two samples only. Next, we compared blood and sperm variants within each individual to assess tissue-specific differences. Approximately 99% of variants were shared between tissues for every individual, with an average of 4,722 unique variants in blood and 4,536 unique variants in sperm (**Supplementary Table S12**). Interestingly, the S1 individual showed a larger number of unique variants, 6,900 in blood and 5,805 in sperm, whereas the other two individuals had fewer: 3,668 and 3,599 unique in blood, and 3,825 and 3,978 unique in sperm, respectively. Notably, unlike the first individual, the latter two showed more unique variants in sperm than in blood.

We next investigated potential mosaic variants characterized by a low VAF (≤ 15%) across all analyzed samples and performed comparative assessments both among different individuals and between tissue types within each individual. We also found high-VAF (>15%) variants that appeared only in blood or only in sperm and also unique to each individual. This pattern suggests that these mutations arose later in development and became restricted to each tissue through separate clonal expansions. Across the three samples, we identified a total of 3,761 mosaic variants in blood tissue, corresponding to an average of 1,278 variants per individual. In sperm tissue, the number of mosaic variants was slightly higher, with a total of 3,881 variants and an average of 1,326 variants per individual (**Supplementary Table S11**). Only eight variants were found to be shared among all samples for blood tissues and four were shared among all individuals of sperm tissues (**Figure 5A&B**). This highlights the expected specificity of mosaic variants. The average VAFs for blood samples ranged from 5.56% to 6.12%, while those for sperm samples were lower and ranged from 4.55% to 5.49% (**Figure 5C and Supplementary Figure S13-16**). The VAF distribution showed that the mean and median VAF of sperm samples were always slightly lower than the blood. Consistent with prior work, tissue-restricted sperm mosaic SNVs often occur at relatively low mosaicism levels, whereas variants detectable in both sperm and blood tend to have higher mosaicism^39^. Interestingly, the mutational signatures of blood and sperm samples of these three individuals showed different patterns among blood and sperm for each individual (**Figure 5D and Supplementary Figure 17-18**). Using a cosine similarity threshold of 0.70 to define a strong match, we identified several sample-tissue combinations that showed high similarity to a set of important COSMIC SBS signatures (**see Supplementary Table S14**). SBS3 exceeded the 0.70 threshold in S1 and in S3 blood, supporting a signal associated with homologous recombination, which is related to DNA repair deficiency. Sample S1 showed a signal (SBS30) consistent with defective base-excision repair (e.g., linked to NTHL1 inactivation). In contrast, Sample S2 did not show any significant COSMIC SBS signatures above this threshold.

**Figure 5:**
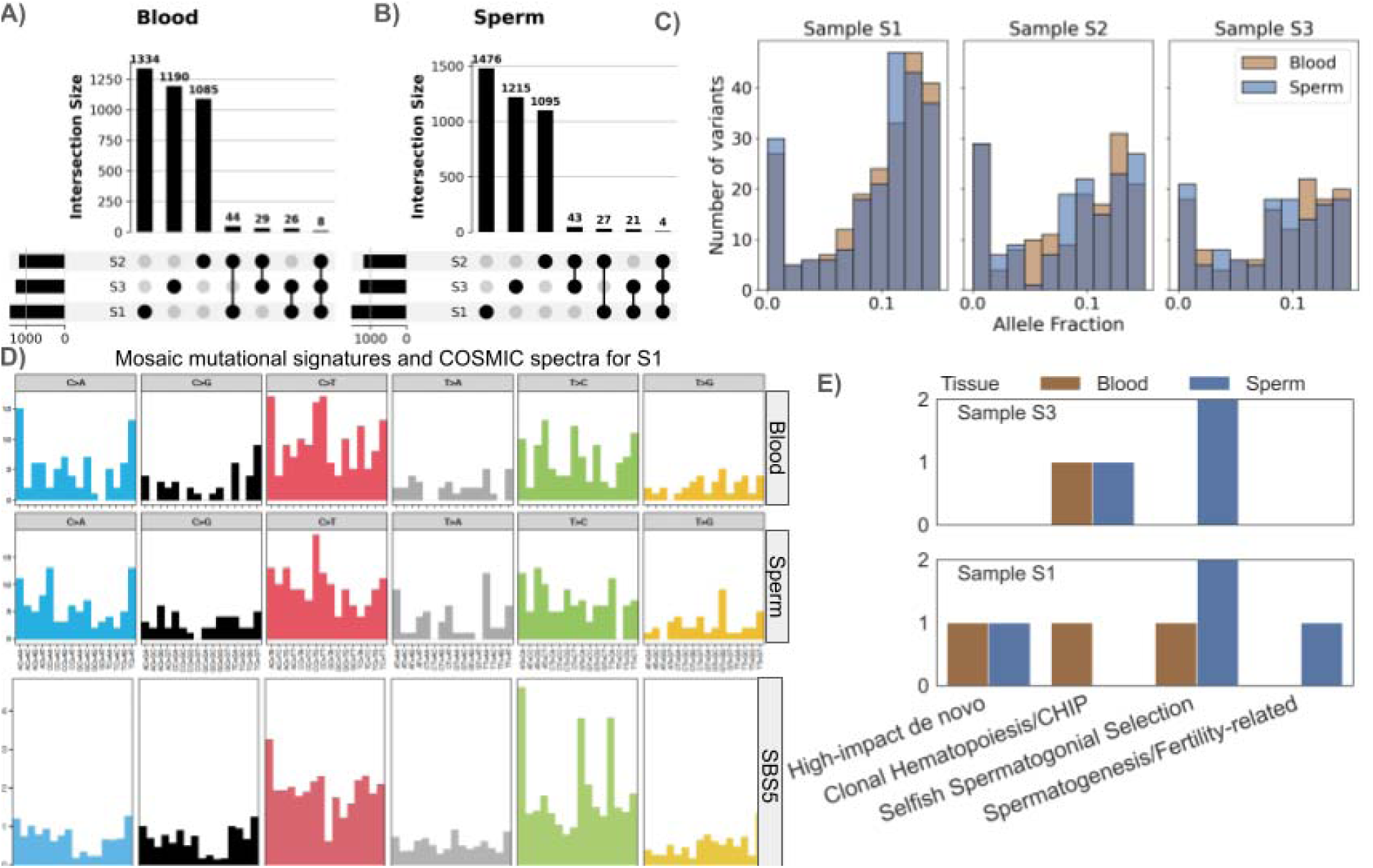
Identification of mosaic variants in fertility related blood and sperm tissues of three individuals. **A)** Shared and unique mosaic variants (VAF <= 0.15) across blood samples of three individuals and **B)** across sperm samples of the same three individuals. **C)** Allele fraction distribution of mosaic SNV and Indels in different blood and sperm tissues of three individuals **D)** Mutational signatures in blood and sperm for sample S1 shows different patterns for blood and sperm for mosaic variants and with SBS5 COSMIC signature (>85% cosine similarity for both tissues) **E)** Clonal dynamics with mosaic variants (known in dbSNP 151) shows blood-only (Hematopoiesis) and sperm-only (Spermatogenesis) variants in hotspot genes for sperm mosaics in Sample S1 and S3.

To systematically assess clonal dynamics of low-VAF mosaic variants, we compiled a curated gene panel for mosaic analysis encompassing four functional categories (see **Supplementary Table S12 and Supplementary Section 2**). Using the above panel of the genes, we identified tissue-specific mosaic variants unique in the blood and sperm samples of the three individuals. The blood-only variants were observed in classical CHIP genes ^40^ with a variant in *JAK2* gene for the first individual and in *TET2 gene* for the third individual. This could reflect age-related clonal hematopoiesis that does not have any transmission risk. Notably, we also detected *JAK2* mosaic variants in the sperm sample of the third individual, and this could suggest occasional clonal expansion in spermatogonia that could confer a potential heritable risk. The sperm-only variants were identified in genes critical for spermatogenesis ^41^, such as one variant in *DMC1* for the first individual, which may affect fertility without necessarily causing developmental disorders. We also identified mosaic variants in paternal age-effect genes^42^, including one variant in *RAF1,* one variant in *FGFR2* for the first individual, and *one in SYCP2, SYCP3,* and *RB1* for the third individual. These mosaic variants potentially reflect selfish spermatogonial expansion and carry a known risk of transmission, often resulting in dominant developmental disorders. Interestingly, we identified two mosaic *SOS1* variants in blood (one UTR3, one intronic), and this likely represents mosaic variants in hematopoietic cells. Finally, variants in the high-impact *de novo* gene *SETD2* were observed in both blood and sperm for the first individual, highlighting loci where germline mosaic variants may contribute to heritable disease (**Figure 5E and Supplementary Table S13**).

Together, these results demonstrate that PAPR, implemented within DRAGEN, enables robust and generalizable detection of mosaic variants in primary tissues without requiring a complete T2T reference. Mosaic variants were largely individual- and tissue-specific, with distinct clonal patterns observed in blood and sperm, including variants affecting genes such as *SETD2* and *JAK2* detected across lineages.

## Discussion

The DRAGEN mosaic low VAF caller establishes a scalable and accurate framework for detecting somatic mosaic single-nucleotide and indel variants from bulk sequencing data, bridging a critical gap between precision and practicality. Through a dedicated machine-learning model integrated directly within the DRAGEN platform, we demonstrate robust sensitivity of 2-3% variant allele fraction (VAF) and above and with appropriate high coverage even down to 1% VAF while maintaining exceptionally low FP rates. Besides the default mode we also introduced a high-sensitivity mode to capture even more mosaic variants, which we only recommend for mosaic variants and not for germline variant calling. Unlike prior approaches, DRAGEN achieves this performance in hours rather than days, enabling high-throughput mosaic analysis across thousands of samples without matched controls. The ability to detect rare mosaic events at population scale redefines what can be achieved with standard Illumina sequencing, transforming low-VAF mosaic variant analysis without matched normal from a specialized challenge into a routine capability for clinical and population genomics.

This work also marks a shift in how mosaic and low-VAF variants can be detected, benchmarked, and validated at scale. With the introduction of the HapMap low-VAF benchmark, we provide the first genome-wide resource for evaluating variant callers in the 1-10% VAF range, which is a regime that has remained largely inaccessible due to the absence of orthogonal benchmark sets. This benchmark bridges the gap between germline and mosaic analyses, offering a unified framework to quantify the sensitivity and precision of low-VAF detection across diverse methods and technologies. We anticipate that this resource will serve as a foundation for community-driven tool development, fostering reproducibility and comparability across future mosaic variant studies.

Beyond accuracy, this work introduces a conceptual shift toward personalized assembly pangenome references (PAPR) for variant discovery. By incorporating phased diploid assemblies into DRAGEN’s pangenome reference, we show that variant calling can be refined in the context of an individual’s own haplotypes while maintaining GRCh38 coordinates for downstream annotation, with benefits on both germline accuracy and mosaic precision. Benchmarking against the Q100 and CMRG datasets revealed consistent gains in both precision and recall for germline variants, particularly in structurally complex and medically relevant regions. The application of this approach to HG002’s blood tissue, the first real-tissue analysis for this benchmark individual, demonstrated that many canonical cell line “mosaic” sites likely represent variants occurring during culture rather than *in vivo*. The benefit of a personalized assembly pangenome reference depends on the quality and completeness of the underlying assembly, with the greatest gains expected in regions where haplotypes are accurately resolved. Overall personalized assembly pangenome references in our cases improved germline variant calling by improving sensitivity and precision. For mosaic calling we did not observe an improvement. This is likely due to the fact that the current HG002 mosaic benchmark is restricted to 85 postulated variants, all of which are identifiable by the default pangenome method. This inherent ceiling in the benchmark set prevents the demonstration of further sensitivity gains with PAPR, as there are no remaining ‘hidden’ variants for the personalized approach to recover. Extending our approach to population-scale pangenomes will enable individualized mosaic discovery at scale, providing a framework for integrating germline, mosaic, and epigenetic variation within a unified genomic coordinate system for which rich annotations are available.

Together, these developments establish DRAGEN as a turning point in mosaic variant analysis. The combination of a publicly available low-VAF benchmark, hardware-accelerated variant calling, and support for personalized assembly pangenome references enables sensitive mosaic variant detection at a scale previously reserved for germline studies. As Illumina sequencing throughput continues to increase, the ability to routinely interrogate low-VAF variants will uncover new biological dimensions ranging from clonal dynamics and tissue mosaicism to early signatures of disease. At the same time, these advances highlight emerging challenges for the field: interpreting the functional relevance of subclonal variants, distinguishing biological mosaic variants from technical artifacts, and defining robust statistical and experimental standards for low-VAF validation. Addressing these challenges will require integrating multi-omic evidence, orthogonal validation, and community-driven benchmarking, which directions are directly enabled by the resources presented here.

Ultimately, this work establishes the foundation for large-scale, tissue-resolved, and longitudinal studies of mosaic genome dynamics. By combining hardware acceleration, pangenome alignment, and a dedicated mosaic ML model, DRAGEN opens the door to the next phase of human genomics, where low-VAF mosaic variants are no longer hidden noise, but a measurable and biologically meaningful component of genome variation.

## Methods

### Human Subjects

Human whole blood and semen were obtained from consenting subjects under Baylor College of Medicine Institutional Review Board Protocol H-12083. All samples were processed and analyzed by the researchers in a de-identified format, with no accompanying personal health information or linkable codes.

### HapMap mix sample

DNA for the three benchmarking samples, HG001 (NA12878, Female), HG002 (NA24385, Male), and HG005 (NA24631, Male), was ordered from Coriell Institute for Medical Research. DNA was initially quantified using a Qubit. Based on this QC, the samples were next diluted to 10 ng/µl, and another round of Qubit was performed on these aliquots for added precision. For three-sample mixes, HG001 and HG002 were first combined in a 2:1 ratio to a total of 1 µg and thoroughly mixed by slow pipetting. From this 2:1 (HG001:HG002) mixture, an additional mixing was performed with HG005 in a 20:1 ratio (HG005: two-sample mix), yielding a final 3 sample mix spike-in ratio of 60:2:1 (HG005: HG001: HG002).

### Illumina sequencing (HGSC)

The HapMap mixed sample consists of HG001, HG002 and HG005 samples from GIAB, mixed *in vitro* before sequencing. The ratio of samples was HG005:HG001:HG002=60:2:1, sequenced at 240x coverage.

Single-cell and bulk samples of the brains were sequenced according to the protocol published here ^21^. The sequencing data were aligned to the reference genome GRCh38 using DRAGEN. The resulting alignments were analysed for coverage using Mosdepth 0.3.10 ^43^.

### PacBio sequencing (HGSC)

Genomic DNA was quantified using the Qubit dsDNA BR Assay Kit (Thermo Fisher Scientific), and DNA size was determined using the Agilent Femto Pulse. Starting with ∼10 µg of DNA, samples were sheared using g-tubes (Covaris Part number 520079) to an average size of 18-22 kb. Sheared DNA was size-selected on the PippinHT instrument (Sage Science) using the 15-20 kb High-Pass definition. Size-selected samples were used as input for the PacBio SMRTBell Prep Kit 3.0 for library preparation. DNA damage repair, A-tailing, and adapter ligation were performed as per the manufacturer’s instructions. The SMRTBell Adapter Index Plate 96A was used to barcode each library. Adapter-ligated DNA was nuclease-treated following the manufacturer’s guidelines and purified using 1X PacBio SMRTBell Cleanup beads. Final libraries were quantified using the Qubit dsDNA quantification High-Sensitivity Assay (Thermo Fisher Scientific). Final library sizes were determined using the Agilent Femto Pulse. Each library was loaded at 325 pM on 2 Revio 25M SMRT Cells and sequenced using SMRTLink v13.1 for a run time of 30 hours.

### DRAGEN variant calling

We used DRAGEN v4.4.6 to produce SNV and Indel callsets, enabling mosaic variant detection. The command below provides an example for the 80× HG002 BAM input.

~~~
/opt/dragen/4.4.6/bin/dragen -f \
--enable-map-align true \
--enable-map-align-output true \
--output-format BAM \
--enable-duplicate-marking true \
--enable-variant-caller true \
--vc-emit-ref-confidence GVCF \
--vc-enable-vcf-output true \
--vc-ml-enable-recalibration true \
--vc-enable-mosaic-detection true \
--vc-mosaic-af-filter-threshold 0.0 \
--enable-cnv false \
--enable-sv false \
--repeat-genotype-enable false \
--enable-vntr false \
--enable-hla false \
--enable-star-allele false \
--enable-targeted false \
--enable-variant-annotation false \
--cnv-enable-self-normalization true \
--logging-to-output-dir true \
--output-file-prefix hapmapmix.80x \
--output-directory <output_dir= \
--bam-input <illumina_80x.bam> \
--ref-dir
~~~

DRAGEN in high-sensitivity mosaic detection mode was run by including `--vc-enable-high-sensitivity-mosaic-detection true` to the above command line.

DRAGEN 4.4.6 runs with default pangenome reference used is hg38-alt_masked.cnv.graph.hla.methyl_cg.rna-11-r5.0-1that is available at the DRAGEN product file page: https://support.illumina.com/sequencing/sequencing_software/dragen-bio-it-platform/product_files.html

### Benchmarking

The benchmarking of HG002 callsets generated from different tools was performed using the GIAB benchmark sets and their corresponding high-confidence regions for SNVs and InDels. The GIAB v4.2.1 SNV dataset was used for genome-wide SNV evaluations, GIAB CMRG v1.0 was used for challenging medically relevant gene regions, and GIAB v1.1 was used for mosaic variant evaluations. The recent release of the GIAB T2T Q100 v1.1-v0.019 benchmark was also used for genomewide SNV benchmarking. The link to the datasets is given below.

GIAB v4.2.1: https://ftp-trace.ncbi.nlm.nih.gov/ReferenceSamples/giab/release/AshkenazimTrio/HG002_NA24385_son/NISTv4.2.1/GRCh38/

GIAB CMRG v1.0: https://ftp-trace.ncbi.nlm.nih.gov/ReferenceSamples/giab/release/AshkenazimTrio/HG002_NA24385_son/CMRG_v1.00/GRCh38/SmallVariant/

GIAB mosaic v1.1 https://ftp-trace.ncbi.nlm.nih.gov/ReferenceSamples/giab/release/AshkenazimTrio/HG002_NA24385_son/mosaic_v1.10/GRCh38/SNV/

GIAB Q100 v1.1-v0.019

https://ftp-trace.ncbi.nlm.nih.gov/ReferenceSamples/giab/data/AshkenazimTrio/analysis/NIST_HG002_DraftBenchmark_defrabbV0.019-20241113/

The benchmarking of HapMap mixture was performed against the benchmark set generated from GIAB SNV v4.2.1 sets of HG001, HG002, and HG005. The benchmark set and the high confidence BED region are available here https://github.com/michalizydo/dragenMosaic.

Data for the YUMC ^14^ benchmark sets were downloaded from PRJNA758606 SRA repository. The data included FASTQ files for cell mixtures of Set-A: M1-1, M1-5, M1-9, M2-9, M2-12, M3-2, M3-12, M3-13.

Per-sample benchmark sets were generated from the positive controls VCF, provided in the original YUCM study. Each variant in this file is annotated by mixture and variant-set labels (e.g., M1:V1, M1:V2, …, M3:V5). Using the design described in Supplementary Table 1 of the original publication ^14^, variants corresponding to the relevant mixture sets were extracted for each sample: M1 samples included variants from sets V1 and V2; M2 samples included V1, V3 and V4; and M3 samples included V1, V3 and V5. Germline and non-variant regions were removed by subtracting coordinates from control VCFs provided within the YUCM study, using bedtools, and the remaining high-confidence regions were additionally restricted to the Agilent V6 exome capture design ^44^.

The evaluations were performed using the vcfeval option of RTGtool ^45^ (v3.12.1) and following is an example of the command and parameters used in our analysis.

~~~
rtgtool vcfeval \
--ref-overlap \
-b <truth_vcf> \
-e <high_confidence_bed> \
-c <callset vcf< \
-t <sdf> \
-o $out \
--sample ALT \
--squash-ploidy \
--vcf-score-field=QUAL \
~~~

In the HG002 mosaic comparisons, additional RTGtool run (parameters as above) was performed on the false-positive (<u>FP.vcf.gz</u>) file from the initial vcfeval, using HG002 Germline benchmark dataset as baseline, to remove any false-positive germline variants, leaving real false-positive mosaics.

### Third-party tools

All benchmark samples have been realigned to the reference genome GRCh38.p14 using BWA-MEM2 ^46^, using `-v 2 -T 20 -K 10000000 -R`

“@RG\\tID:lib1\\tLB:lib1\\tSM:$<sample>\\tPL:illumina\\tPU:unit1”

parameters to ensure read group identification and reproducibility. The .bam files were split per-chromosome and in case of DeepMosaic ^12^ and MosaicForecast ^11^ variants were called on each using Mutect2 ^11,13^:

`Mutect2 -R grch38_genome_mainchrs_bwa.fa -I <input> -O <output>`.

DeepSomatic internal pipeline includes proprietary variant calling - the entire pipeline was run according to GitHub instructions for tumor-only mode ^17^.

For DeepMosaic, bam files containing only primary alignments were generated. DeepMosaic pipeline was run according to GitHub manual ^12^, with feature extraction and mosaicism prediction steps performed independently per-chromosome. Finally, a custom script was used to copy DeepMosaic result as an annotation to each of the original VCF-files, which were concatenated and filtered for calls containing annotations.

For MosaicForecast, the pipeline was run according to GitHub instructions ^11^. Read-level feature extraction and genotype prediction steps were performed independently per-chromosome.

Custom script was used to copy genotype prediction into original VCF as a tag and non-classified/malformed tags were removed.

All custom scripts and step-by-step instructions are available on Github: https://github.com/michalizydo/dragenMosaic.

### Benchmarking set for HapMap mixed sample

The mosaic SNV benchmark set for the HapMap mixed samples (HG001, HG002, and HG005) was generated by combining the Genome in a Bottle (GIAB) SNV benchmark sets (GRCh38, v4.2.1) for these samples. The VCF-file and BED regions were downloaded from the following links:

HG001: https://ftp-trace.ncbi.nlm.nih.gov/ReferenceSamples/giab/release/NA12878_HG001/NISTv4.2.1/GRCh38/

HG002:https://ftp-trace.ncbi.nlm.nih.gov/ReferenceSamples/giab/release/AshkenazimTrio/HG002_NA24385_son/NISTv4.2.1/GRCh38/

HG005:https://ftp-trace.ncbi.nlm.nih.gov/ReferenceSamples/giab/release/ChineseTrio/HG005_NA24631_son/NISTv4.2.1/GRCh38/

The goal was to identify variants present in HG001 and HG002 but absent in HG005. To begin, the high-confidence BED regions of all three samples were intersected using bedtools (v2.30) to define genomic regions shared across the samples. The SNV benchmark VCF files for HG001, HG002, and HG005 were then merged with bcftools (v1.19) merge. Bi-allelic variants (-m2 -M2) and multi-allelic variants were separated into distinct VCF-files. To simplify the benchmark, multi-allelic sites introduced during merging were excluded, and their corresponding regions were also removed from the intersected BED file. The candidate somatic mosaic variants were filtered from the bi-allelic variant file and defined as those not observed as germline in HG005. Finally, BED regions containing HG005 germline variants were filtered out from the high-confidence regions. The complete scripts to generate the benchmark set and the high-confidence region are available here: https://github.com/michalizydo/dragenMosaic.

### Identifying mosaic variants

The DRAGEN (v4.4.6/v4.3.6) pipeline was run in mosaic mode to call variants. The variants located on autosomes within the GIAB high-confidence regions, passing the **PASS** filter and having a variant allele fraction (VAF) ≤ 15%, were classified as mosaic calls. The following bcftools command was used to generate mosaic calls.

`bcftools view -t chr1-22 -f PASS -R GIAB.bed -i “FMT/AF<=0.15” -Oz -o mosaic.vcf.gz input.vcf.gz`

### Brain analyses

Bulk brain DRAGEN calls were merged with single-cell DRAGEN calls using bcftools^47^ merge command:

`bcftools merge -Oz -m all -o <output> --threads 24 --force-samples <input>`

Mosaic variants are separated from germline by bcftools view -i “FMT/AF<=0.15” command Custom scripts are used to summarize the overlap of variants between bulk sequencing and corresponding single cells, as well as VAF distribution and heatmap.

To generate mutational spectra across the bulk brain samples, spectra.R script was used, accompanied by cosine.R that show cosine similarity heatmap between samples. Per-brain hotspots of mutational load and their overlap with exon locations extracted from UCSC table browser^48^ for GRCh38/hg38 reference genome build were performed with custom scripts available on the Github. The gene names at hotspots were used as input for GOrilla gene ontology analysis ^49^.

### Building a personal assembly pangenome reference

The haplotypes assemblies of HG002 Q100 v1.1 from T2T-consortium and HPRC^37^ were used to build a personalized assembly pangenome reference. The DRAGEN(v4.3.6) pangenome VCF builder app (https://supportassets.illumina.com/products/by-type/informatics-products/basespace-sequence-hub/apps/dragen-pangenome-vcf-builder.html) used the haplotype-resolved assemblies, i.e., hap1 and hap2 fasta files for the sample and GRCh38 reference to generate a VCF file HG002 containing the SNV calls. Then, the VCF file and metadata CSV that includes the sample name i.e., HG002, fasta file paths, sample ID, cohort info, etc. were provided to DRAGEN’s multigenome reference builder app (https://www.illumina.com/products/by-type/informatics-products/basespace-sequence-hub/apps/dragen-multigenome-reference-builder.html) to generate a pangenome reference hash table.

### DRAGEN Mosaic detection caller

The DRAGEN Mosaic detection workflow described in **Figure 1A**, leverages the highly-optimized infrastructure and advanced algorithms used by the DRAGEN germline workflow described in detail in^16^. The DRAGEN mosaic detection algorithm exploits the DRAGEN pangenome reference to recover low variant allele fraction (VAF) calls, without requiring matched controls. Calls made by the mosaic detection algorithm are integrated into a standard VCF output alongside germline variants, and tagged to ease interpretation.

To place the DRAGEN Mosaic detection algorithm into context, we briefly summarize the DRAGEN germline workflow. Given an input set of reads for the sample, e.g., HG002, and a pangenome reference genome build on a target reference genome, e.g., GRCh38, the reads are mapped and aligned to the pangenome reference using the DRAGEN multi-genome mapper, and reported in the target reference genome coordinates. After being sorted, the aligned reads are streamed to the germline variant caller that identifies sufficient coverage and evidence of variants in the reads to establish active regions. The reads covering an active region are locally assembled via a de Bruijn graph to generate a set of candidate haplotypes. The HMM then computes a likelihood for each read-haplotype pair and candidate genotypes are formed from diploid combinations of variant events (SNV or indel). Genotyped candidates are then presented to DRAGEN-ML that refines the genotyped candidates in a computationally efficient manner.

The DRAGEN mosaic detection algorithm, when enabled, enhances the DRAGEN germline workflow by attempting to rescue genotyped candidates, usually with low allele fraction, that are rejected by the germline DRAGEN-ML model. This task is performed by presenting the rejected candidate events to an auxiliary Machine Learning (ML) model specialized in capturing signatures proper of somatic mosaic variants. Genotyped candidates rescued by the mosaic ML model are emitted to the VCF file with genotype set to 0/1 and tagged with a INFO/MOSAIC tag indicating that the variant was called by the mosaic detection algorithm.

The DRAGEN mosaic detection workflow, consisting of the DRAGEN germline workflow with mosaic detection enabled, achieves remarkable recall and accuracy through three key enhancements: (1) We improve sensitivity by recovering reads in low-mappability regions using the DRAGEN pangenome reference and associated advanced alignment algorithms ^16^ (2) We extract active regions with a lower evidence threshold. This allows positions with lower read evidence to progress through the pipeline increasing sensitivity. (3) We use a ML model that is trained specifically to identify somatic mosaic variants improving specificity.

We further extend the DRAGEN mosaic detection workflow with a high-sensitivity mosaic detection mode, designed to increase the sensitivity detection of low-AF variants at the expense of a moderate increase in variant calling runtime. This is achieved by further lowering the evidence threshold for active region and candidate events generation to allow more candidate events through the variant calling and genotyping pipeline, as well as expanding the model training dataset to include lower-AF variants and in-silico admixture datasets at high depth coverage.

The mosaic model, in both regular and high-sensitivity mosaic detection modes, is trained using a supervised learning approach. Due to the lack of validated somatic mosaic datasets, we generated synthetic ones using Bamsurgeon ^50^ to simulate SNV and Indel somatic mosaic variants into germline datasets. We edited both WGS and WES datasets from the GIAB reference samples, introducing ∼50k and ∼10k mosaic variants, respectively. The variants were introduced in reference-homozygous positions identified in the GIAB v4.2.1 high-confidence bed, with variant allele fractions uniformly distributed between 1% and 45%. We simulate mosaic variants in a range of sequencing platforms and configurations to allow the trained model to generalize across different sequencers, depths, lab-preparation flows, coverages, etc. Because real mosaic variants present in the underlying samples are not represented in the synthetic benchmark set, some of these true variants initially appear in the negative training pool affecting model performance. To address this label-noise issue, we applied a negative filtering procedure. A preliminary model was trained on the initial dataset, then used to score all negative sites. Negatives with high predicted mosaic probability were flagged as potential true positives and removed from the negative set. The model was retrained on the refined dataset which is a cleaner and more accurate training set. Finally, the model is tested using validated somatic mosaic datasets, *in silico* admixture datasets, and reference datasets.

The mosaic model is trained using rich read-level features including statistical descriptions of mapping quality, base quality, strand bias, variant length, GC bias, depth, AF, as well as internal HMM scores, including foreign read probabilities, SSE triggers, base quality, and other statistics from VC internal processing. These features are extracted during DRAGEN variant calling. The features are used to build a model using offline training, outside the DRAGEN pipeline. The model uses a gradient-boosted ensemble of weak decision tree learners to identify mosaic variants, resulting in a very efficient and accurate model (adds only a few minutes to variant calling time without requiring hardware acceleration).

The data we use for training do not attempt to model real somatic mosaic data prior probabilities. Therefore, in contrast to the gemline ML model, the mosaic ML prediction probabilities may result being not well calibrated - the ML model output probability represents relative confidence in the call. Mosaic variants are output in the same VCF file as germline variants. For a called mosaic variant, the QUAL field is then updated with a confidence score calculated from the model probability output. The mosaic pass threshold calibration of the QUAL field to recover high-confidence mosaic events based on validated mosaic data.

## Data availability

GIAB SNV germline benchmark VCFs (v4.2.1) and high-confidence bed regions are available in the following links:

- HG001: https://ftp-trace.ncbi.nlm.nih.gov/ReferenceSamples/giab/release/NA12878_HG001/NISTv4.2.1/GRCh38/
- HG002:https://ftp-trace.ncbi.nlm.nih.gov/ReferenceSamples/giab/release/AshkenazimTrio/HG002_NA24385_son/NISTv4.2.1/GRCh38/
- HG005:https://ftp-trace.ncbi.nlm.nih.gov/ReferenceSamples/giab/release/ChineseTrio/HG005_NA24631_son/NISTv4.2.1/GRCh38/

GIAB Challenging Medically Relevant Gene (CMRG) benchmark v1.00 is available here: https://ftp-trace.ncbi.nlm.nih.gov/ReferenceSamples/giab/release/AshkenazimTrio/HG002_NA24385_son/CMRG_v1.00/GRCh38/SmallVariant/

GIAB T2T Q100 v1.1 benchmark is available here: https://ftp-trace.ncbi.nlm.nih.gov/ReferenceSamples/giab/data/AshkenazimTrio/analysis/NIST_HG002_DraftBenchmark_defrabbV0.019-20241113/

GIAB mosaic benchmark v1.1 is available here https://ftp-trace.ncbi.nlm.nih.gov/ReferenceSamples/giab/release/AshkenazimTrio/HG002_NA24385_son/mosaic_v1.10/GRCh38/SNV/

The benchmark VCF and high confidence BED regions are available here: https://doi.org/10.5281/zenodo.18200155

HG002 assemblies used for personalized pangenome: https://github.com/human-pangenomics/hprc_intermediate_assembly/blob/main/data_tables/assemblies_release2_v1.0.index.csv

DRAGEN 4.4.6 default pangenome reference hg38-alt_masked.cnv.graph.hla.methyl_cg.rna-11-r5.0-1: https://s3.us-east-1.amazonaws.com/webdata.illumina.com/downloads/software/dragen/references/genome-files/hg38-alt_masked.cnv.graph.hla.methyl_cg.rna-11-r5.0-1.tar.gz

Yonsei University College of Medicine (YUCM) benchmark data is available at PRJNA758606 SRA/Bioproject 758606 repository:

https://www.ncbi.nlm.nih.gov/bioproject/758606

Information from the original publication^14^ was used in conjunction with the provided variant set to construct the truthset:

https://github.com/hiyoothere/Benchmarking-mosaic-variant-detection/blob/master/controls/Positive_controls_hc.vcf

Germline and non-variant regions were removed by subtracting coordinates from control VCFs provided within the YUCM study, using bedtools, and the remaining high-confidence regions were additionally restricted to the Agilent V6 exome capture design ^44^:

https://github.com/hiyoothere/Benchmarking-mosaic-variant-detection/blob/master/controls/Negative_controls_nonvariant.vcf.gz

https://github.com/hiyoothere/Benchmarking-mosaic-variant-detection/blob/master/controls/Negative_controls_germline.vcf

https://raw.githubusercontent.com/AstraZeneca-NGS/reference_data/refs/heads/master/hg38/bed/Exome-Agilent_V6.bed

Illumina bulk WGS from MSA brains have been deposited in dbGaP (accession #: phs001963.v3.p1) under the genome IDs MSA00915 (for MSA1_CingCx) and MSA00911 (for MSA2_CingCx). Single-cell sequence data has been deposited at the European Genome-phenome Archive (EGA), which is hosted by the EBI and the CRG, under accession number EGAS50000000020 (Dataset ID: EGAD50000000030)

Benchmark for HapmapMix: https://github.com/michalizydo/dragenMosaic.

## Code availability

All scripts for the analysis and benchmark set generation are available here: https://github.com/michalizydo/dragenMosaic.

DRAGEN v.4.4.6 is available as a free trial for academic institutions upon request at and is hosted on AWS.

## Acknowledgements

FJS, MBI, SB, TG are partially supported by NIH grants (1U01HG011758, 4UH3NS132105, 1UM1DA058229, R01HD106056). T.M.N. was supported by a Medical Scientist Training Program grant from the National Institute of General Medical Sciences of the National Institutes of Health under award number: T32GM152349 to the Weill Cornell/Rockefeller/Sloan Kettering Tri-Institutional MD-PhD Program. OS.R-A. was supported by the Cornell University Graduate School Deans Excellence Fellowship. This research was supported in part by the Intramural Research Program of the National Institutes of Health (NIH) (program #: ZIANS003154). The contributions of the NIH author SWS were made as part of their official duties as NIH federal employee, are in compliance with agency policy requirements, and are considered works of the United States Government. However, the findings and conclusions presented in this paper are those of the author and do not necessarily reflect the views of the NIH or the U.S. Department of Health and Human Services. CP acknowledges support by the MSA Trust. MSA brain tissue samples were provided by the UCL Queen Square Brain Bank for neurological disorders. We are grateful to the individuals who donated their brains.

## Contributions

SB, MR, YW, MBI, GP, CM, CP, JH, RM, SC, and FJS designed the study. SB, MR, YW, MBI and DT performed *in-silico* experiments and analyses. SB, MR, YW, MBI, and AV were involved in method development. CLD, EK-E, KK, HM, SWS, HS, XZ, HD, CM, TG collected and provided experimental data/performed experiments. TMN and OSR-A managed data. SB, MR, YW, MBI, CP, FJS wrote the manuscript text. All authors contributed to manuscript review.

