## Supplement for "Scalable and comprehensive mosaic variant calling using DRAGEN"

#### A)

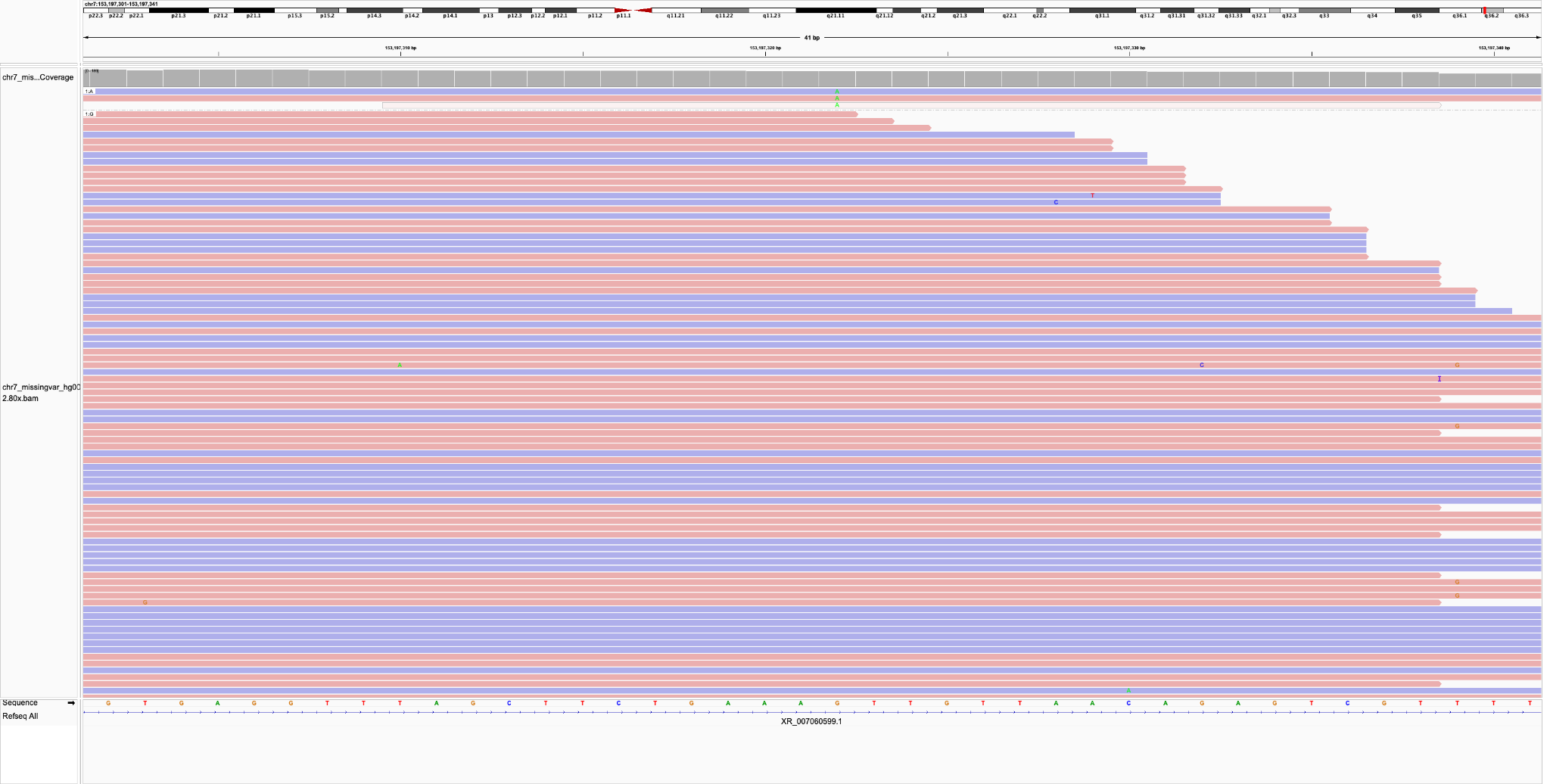

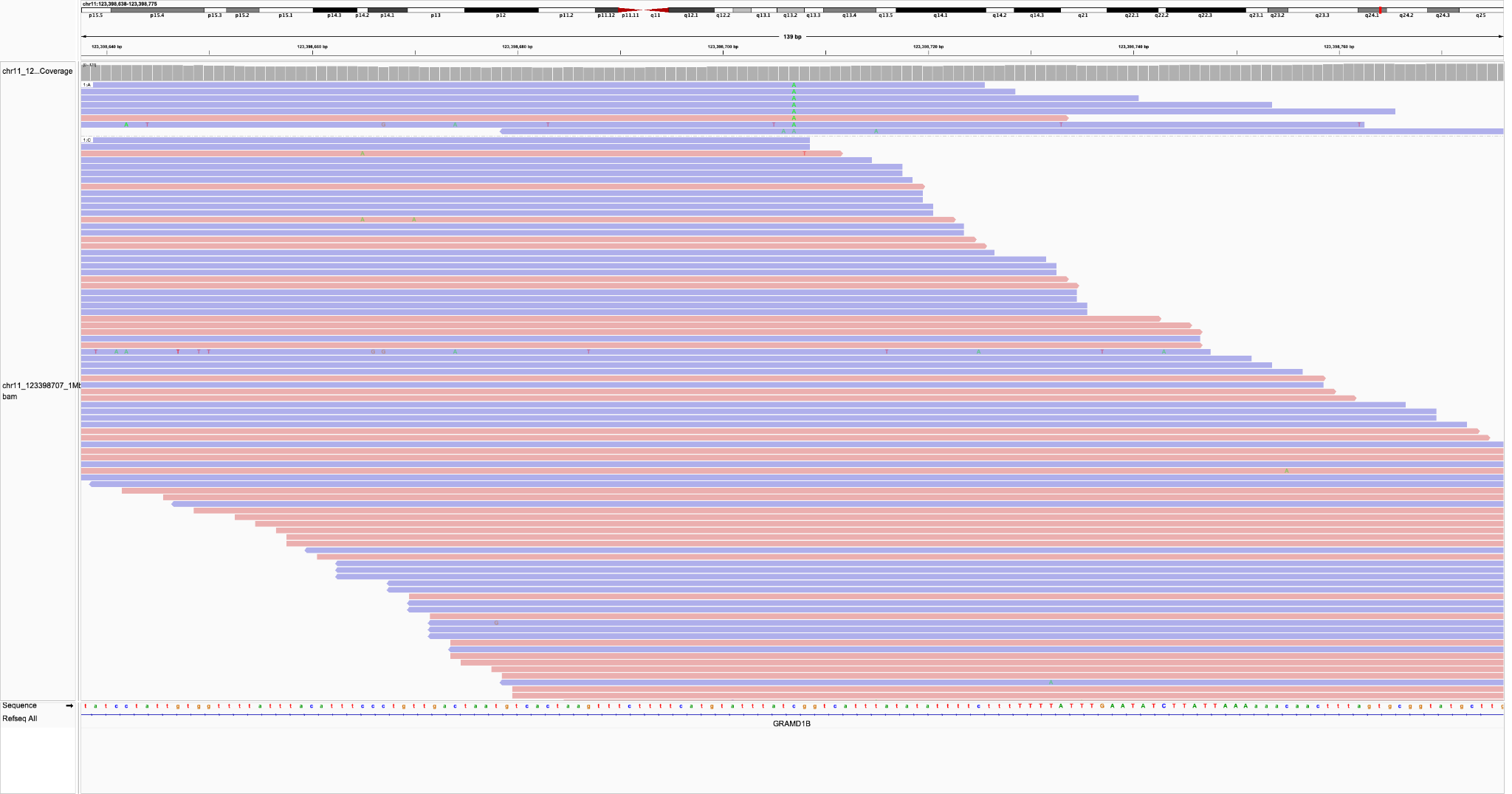

B)

***Supplementary Figure S1****: HG002 GIAB small-variant benchmark putative downsampling artifacts - the top missing benchmark variant is present only on 2 reads after downsampling (third read with MQ=0); the bottom missing variant AF drops from 11% in the 500x sample to 7% in the 80x sample; the downsampled pileup has high strand bias as 7 out of 8 reads are aligned to the reverse strand, while in the non-downsampled pileup we have 21 forward and 30 reverse aligned reads; the non downsampled pileup has 51 supporting reads with 44 with “F” base quality, 4 with “:” base quality and 3 with “,” base quality, while the downsampled pileup has 5 with “F” base quality, 2 with “:” base quality, and 1 with “,” base quality.*

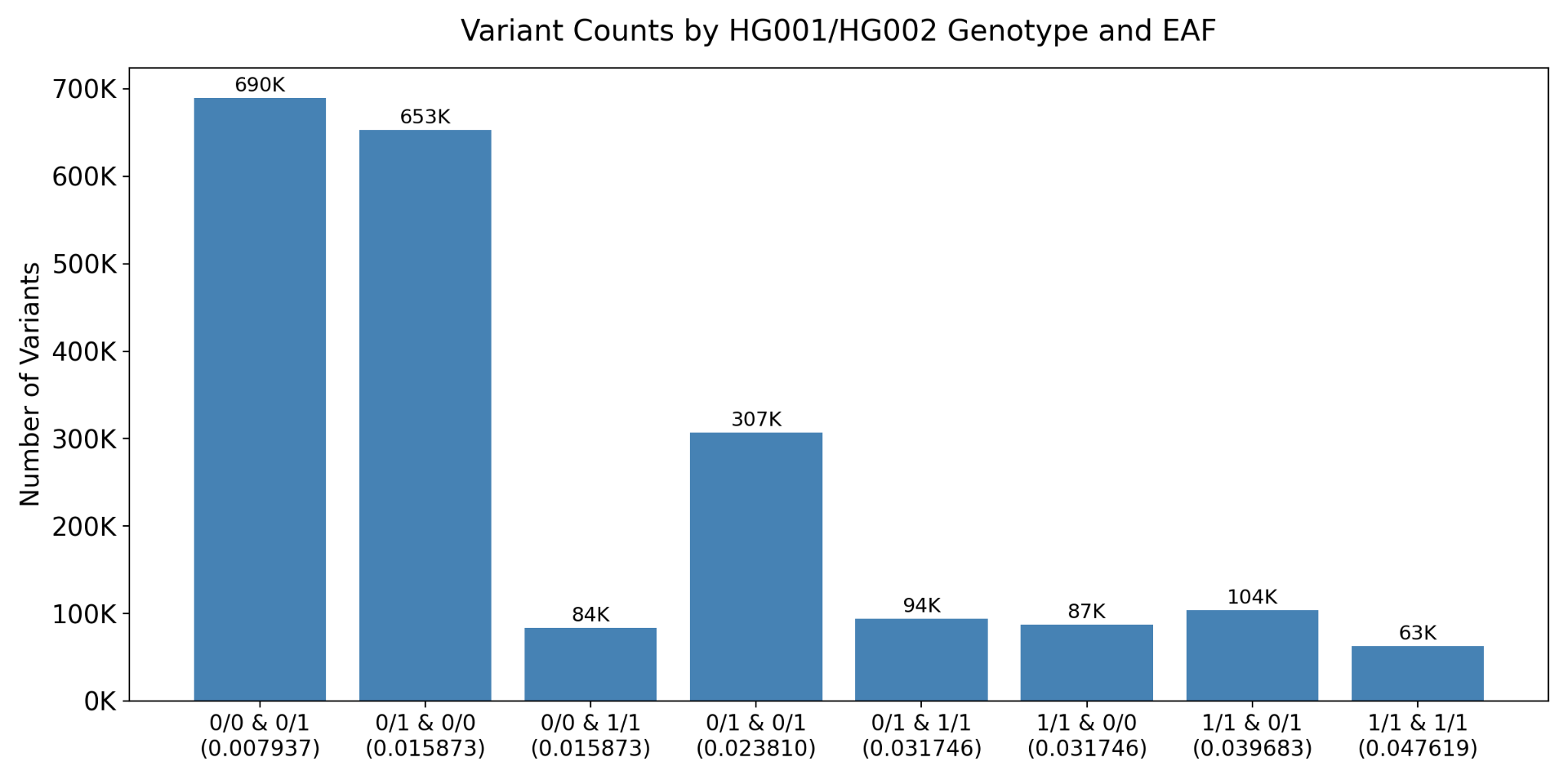

***Supplementary Figure S2:*** *The distribution of benchmark variants in the benchmark set based on the HG001 and HG002 GT combinations.*

#### Benchmarking of HapMap callsets of DRAGEN

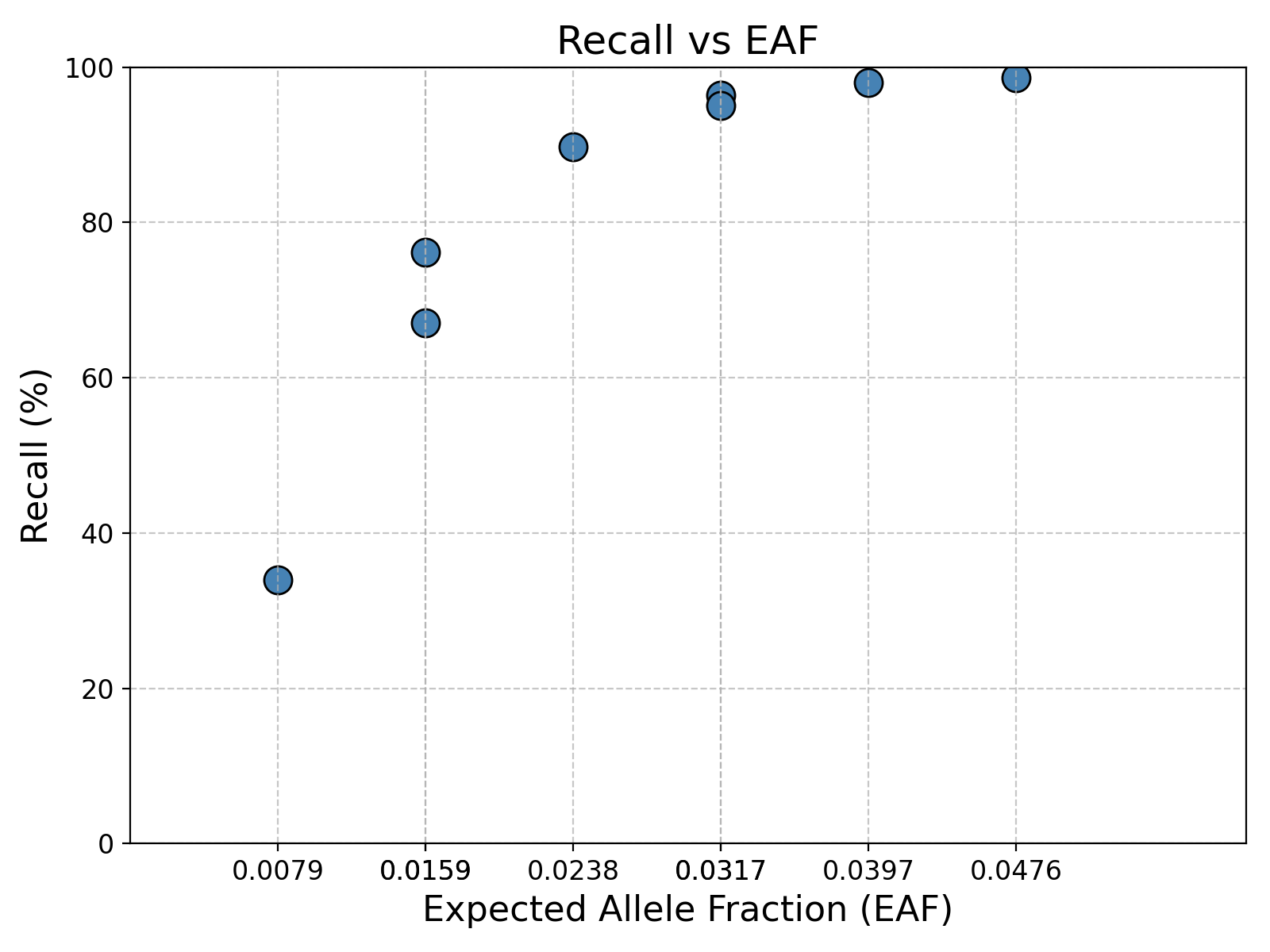

***Supplementary Figure S3:*** *The evaluations of sensitivity* *for* *DRAGEN mosaic calls on the HapMap mix 240x sample based on expected VAF bins due to different GT combinations of HG001 and HG002.*

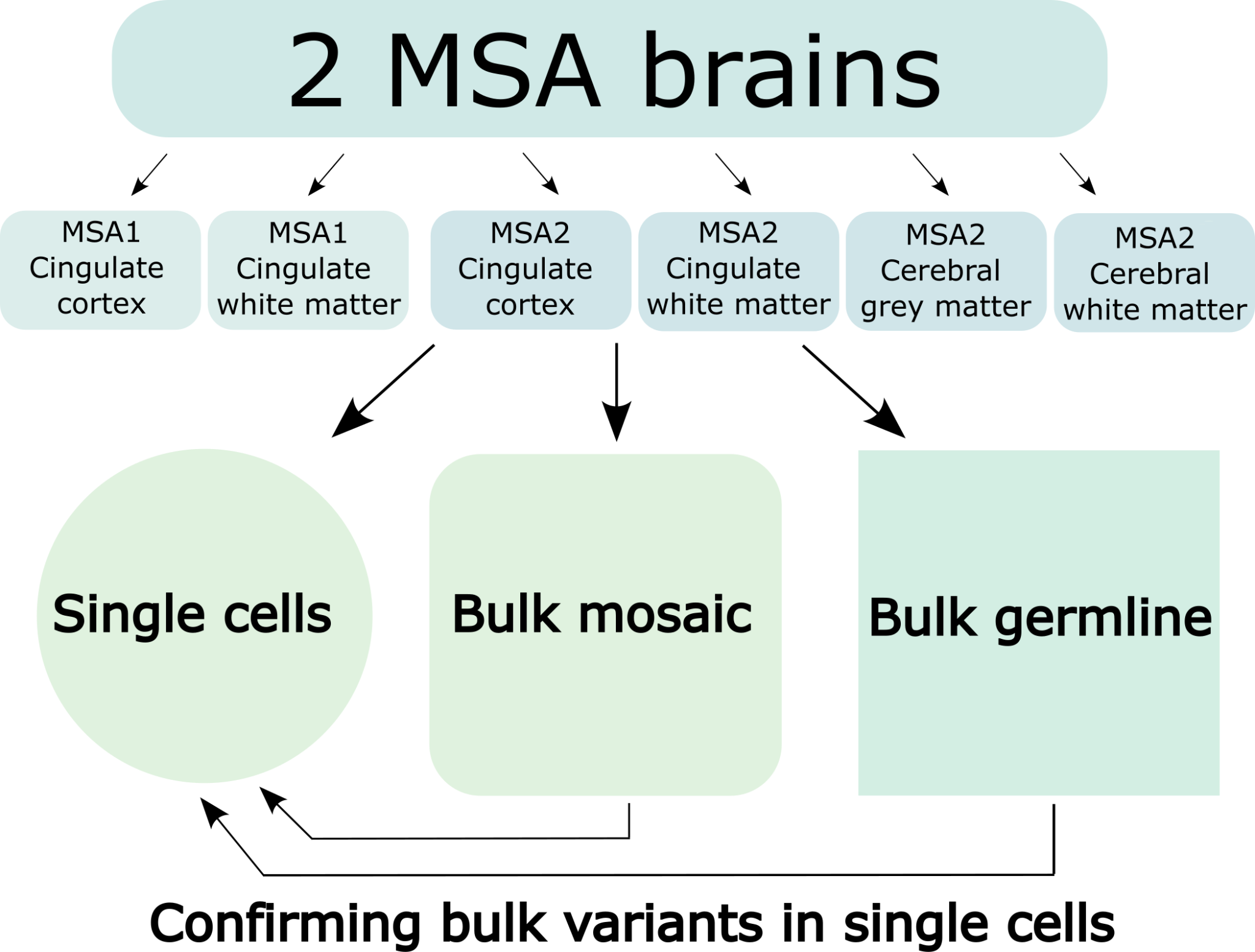

***Supplementary Figure S4***: *Schematic of variant comparisons in the brains.*

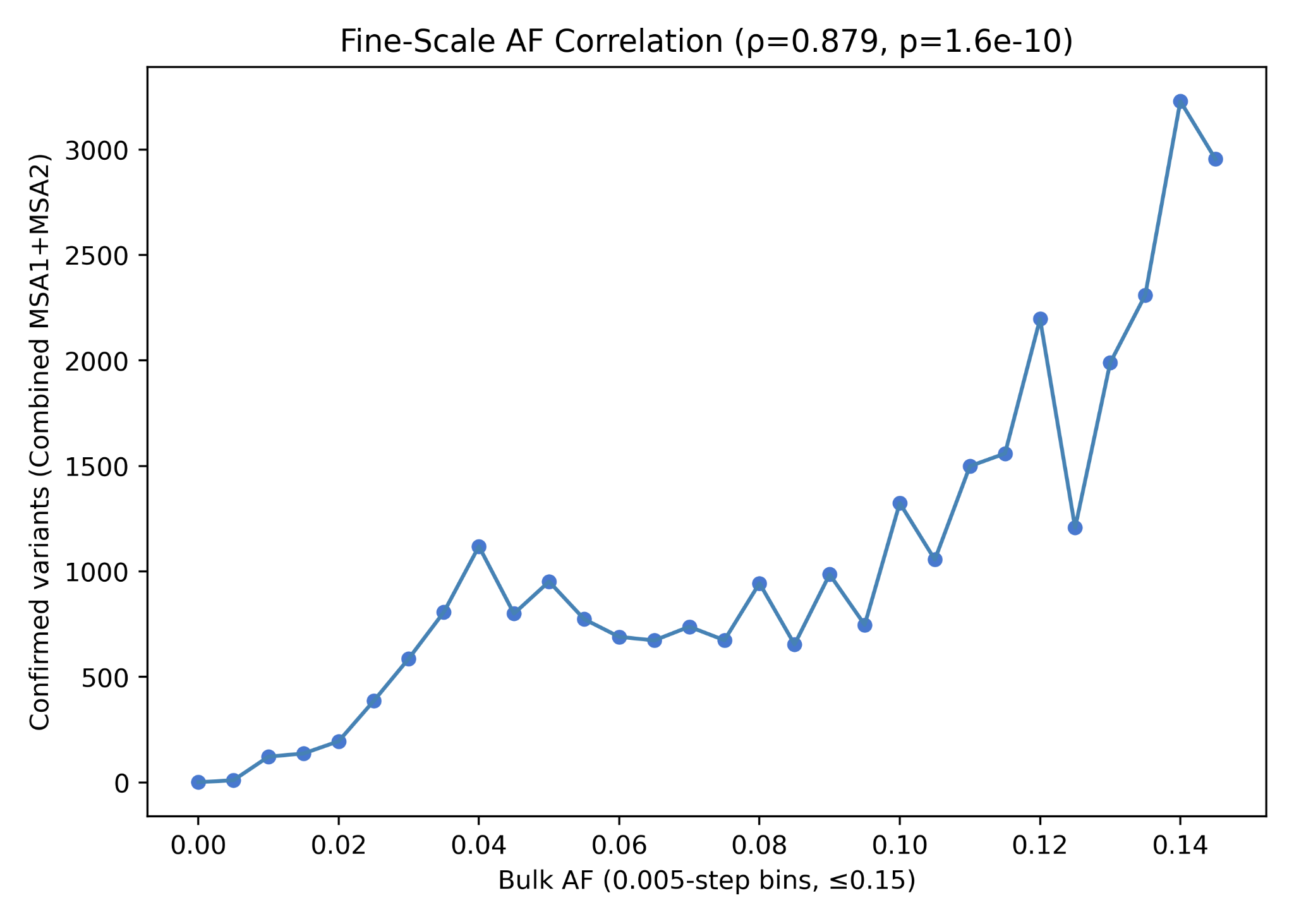

***Supplementary Figure S5****:* *Variant Allele Fraction (VAF) spectrum of bulk brain mosaic variants confirmed in corresponding single cells.*

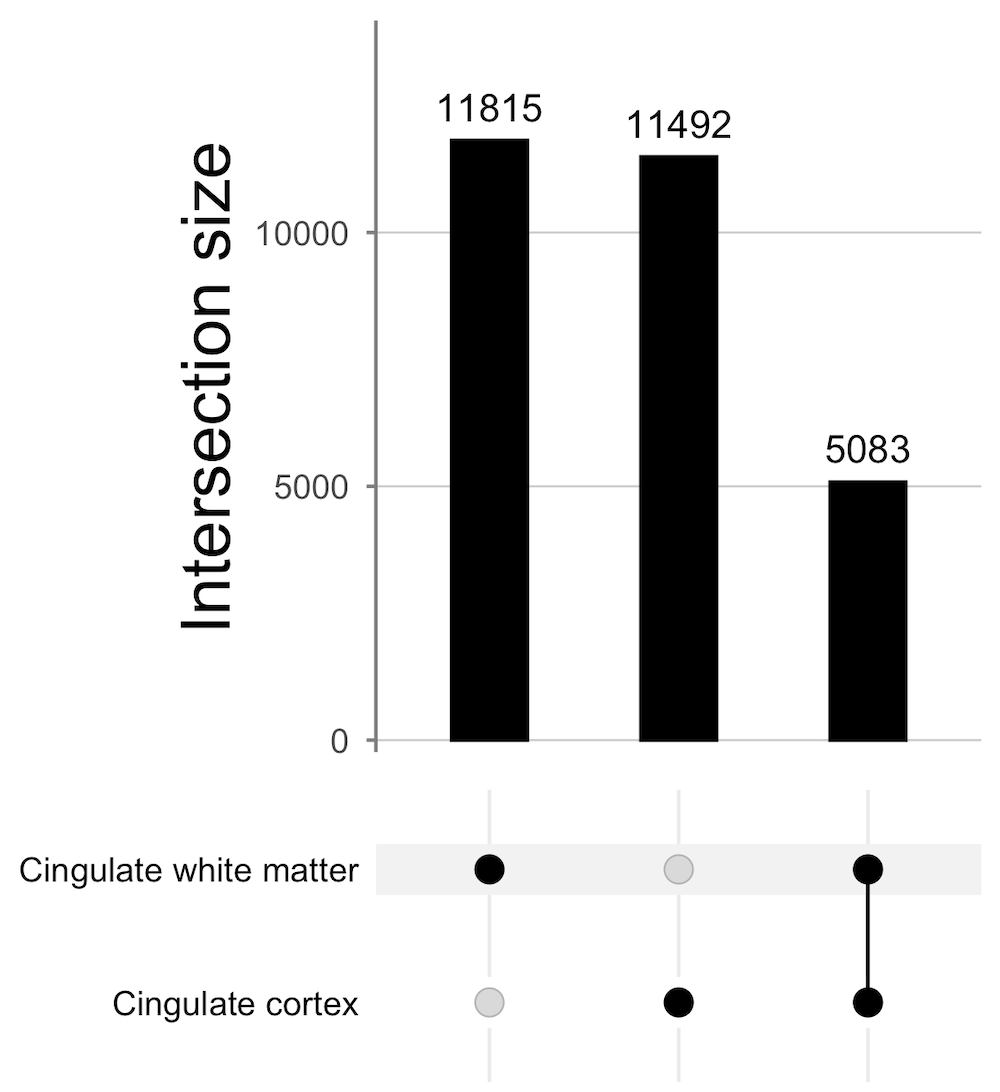

***Supplementary Figure S6***: *Overlap between variant counts in MSA1 brain samples - Cingulate cortex and cingulate white matter regions.*

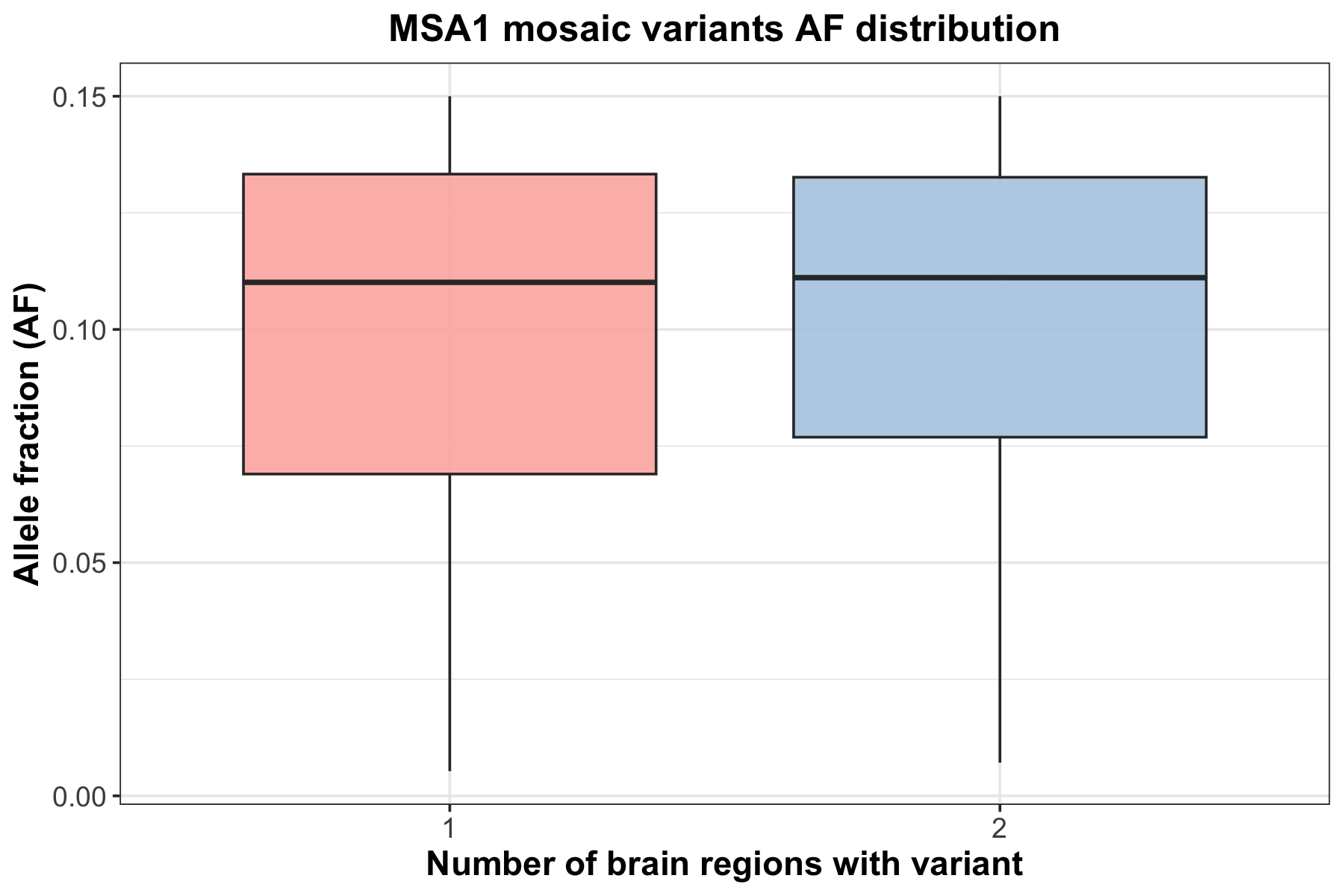

***Supplementary Figure S7***: *Mosaic variant AF distribution and its presence in MSA1 brain.*

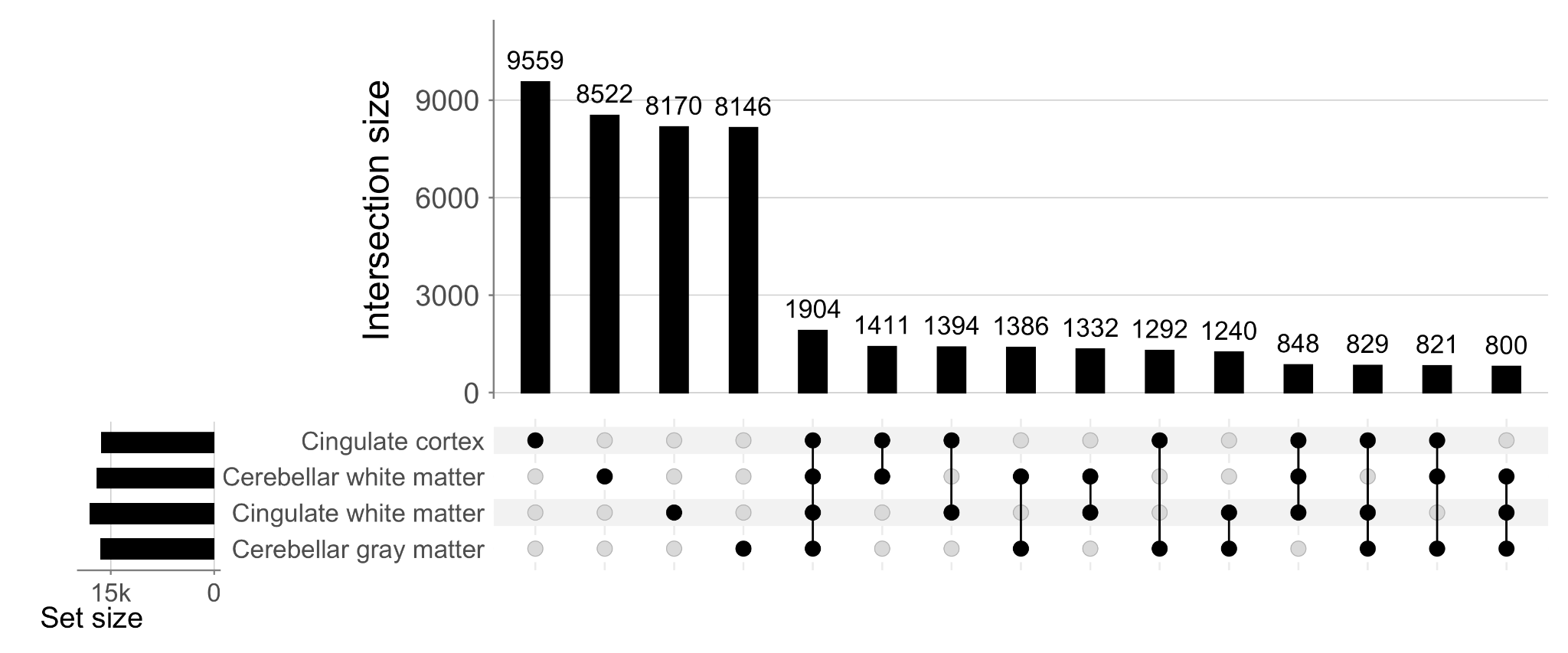

***Supplementary Figure S8***: *Overlap between variant counts in MSA2 brain samples - Cingulate cortex, cingulate white matter, cerebellar white matter and cerebellar gray matter regions.*

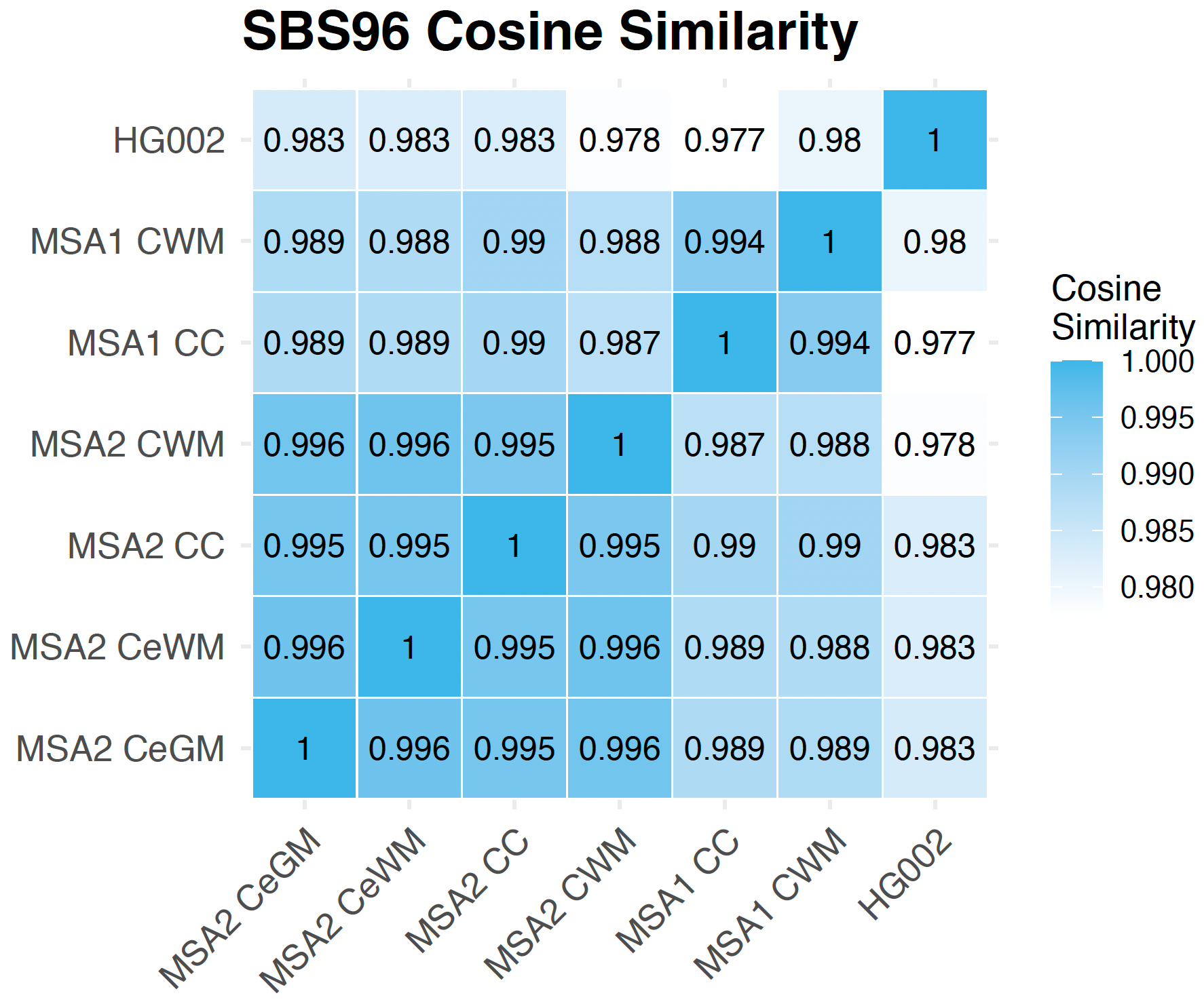

***Supplementary Figure S9*:** *SBS96 Cosine similarity matrix between tested brain regions and HG002, for all mosaic variants (AF<=0.15).*

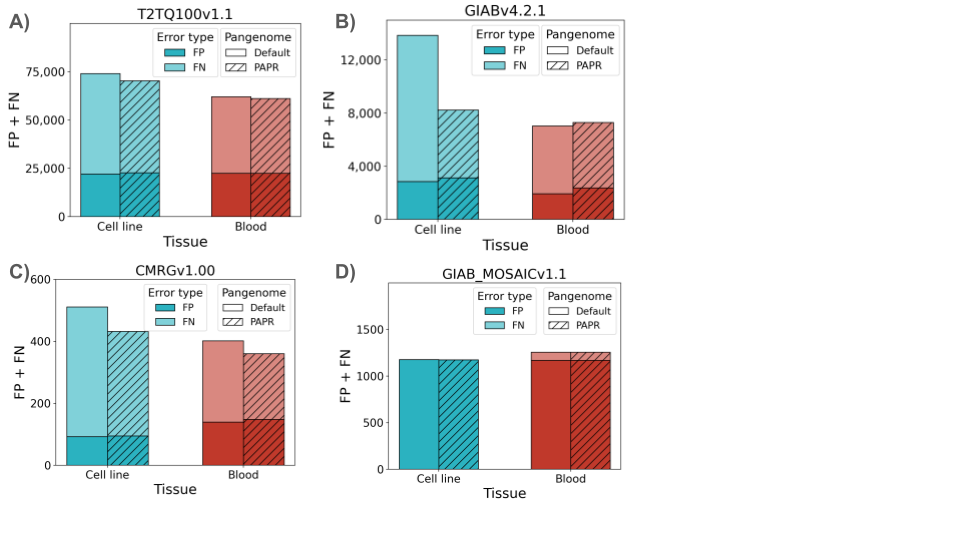

***Supplementary Figure S10:*** *Total errors (False positive + False negatives) of DRAGEN calls with default and PAPR pangenome for HG002 cell line and blood tissues* ***A)*** *benchmarking against the GIAB T2T v1.1 genomewide SNV and Indel benchmark set* ***B)*** *benchmarking against the GIAB v4.2.1 benchmark set* ***C)*** *benchmarking against the GIAB CMRG v1.00 and* ***D)*** *benchmarking against the GIAB mosaic v1.1*

### A)

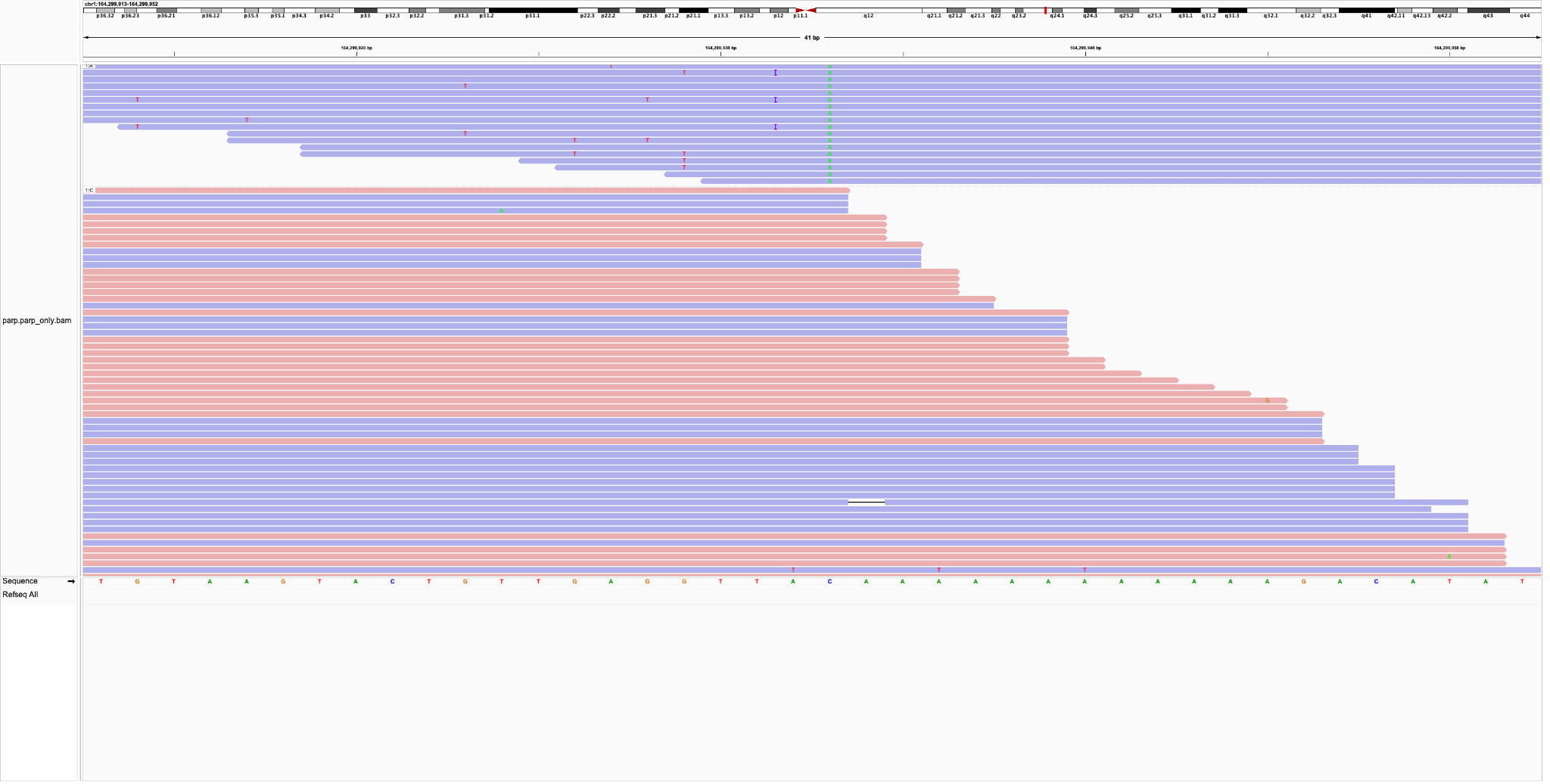

B)
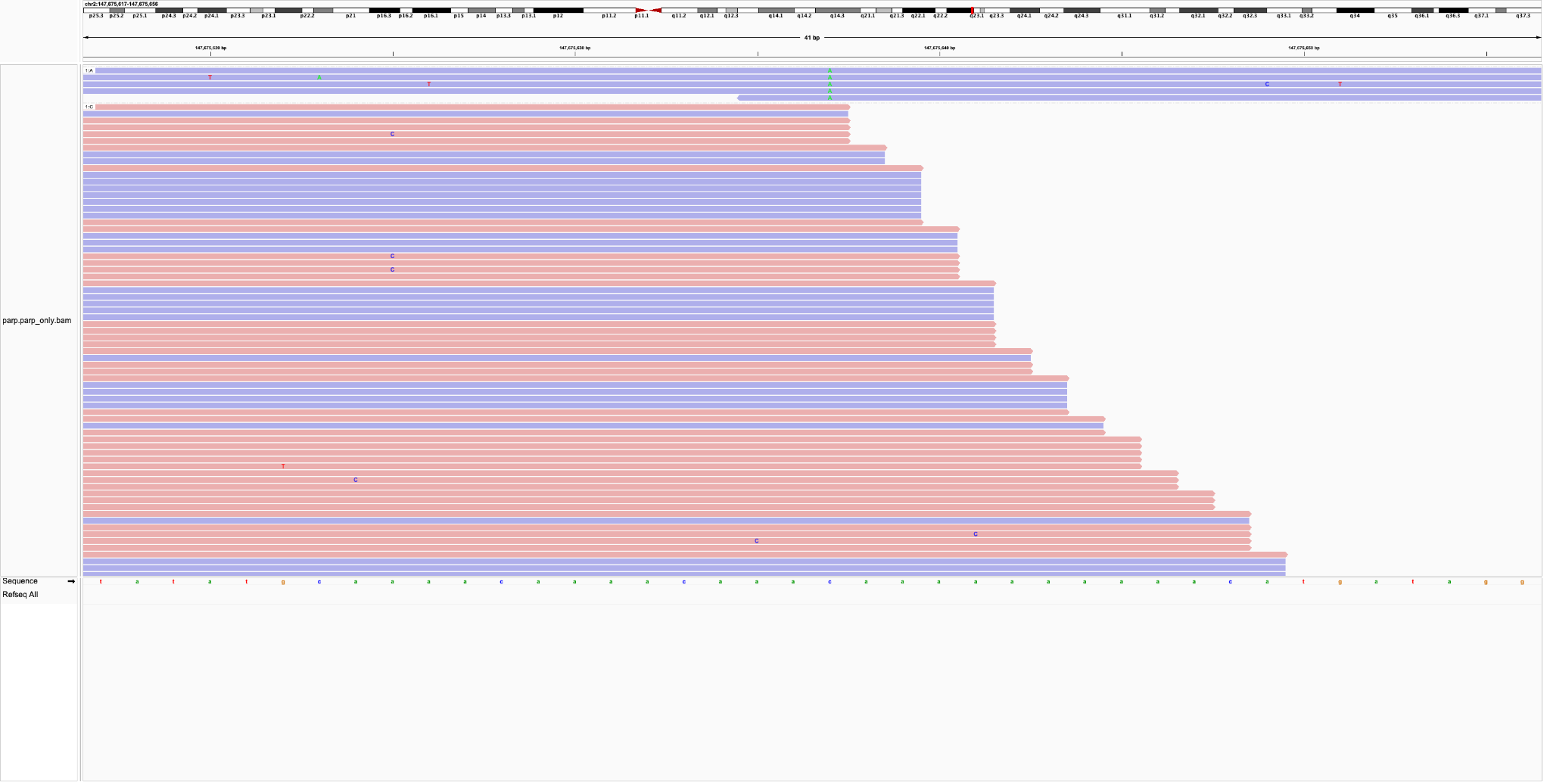

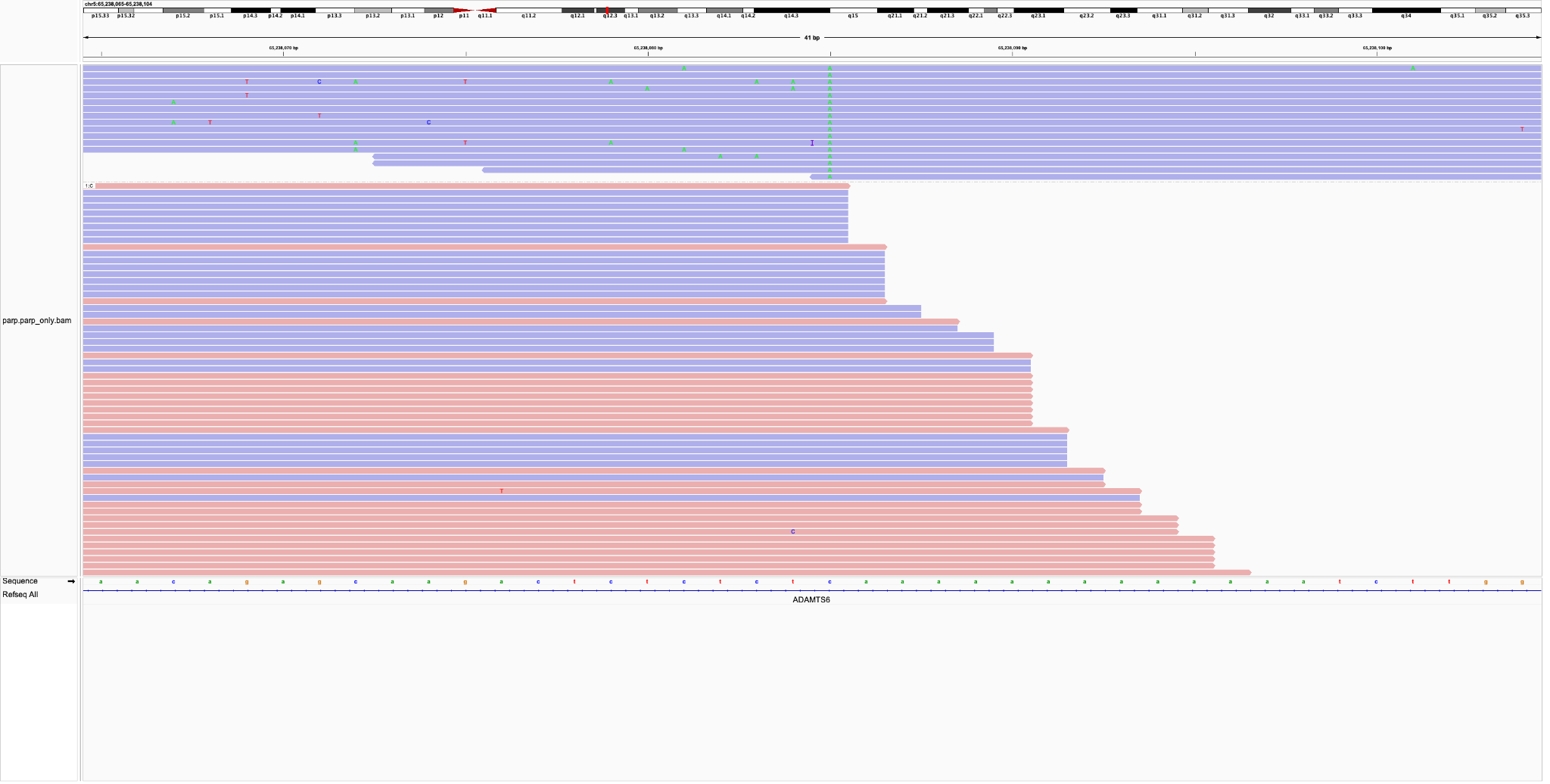

C)

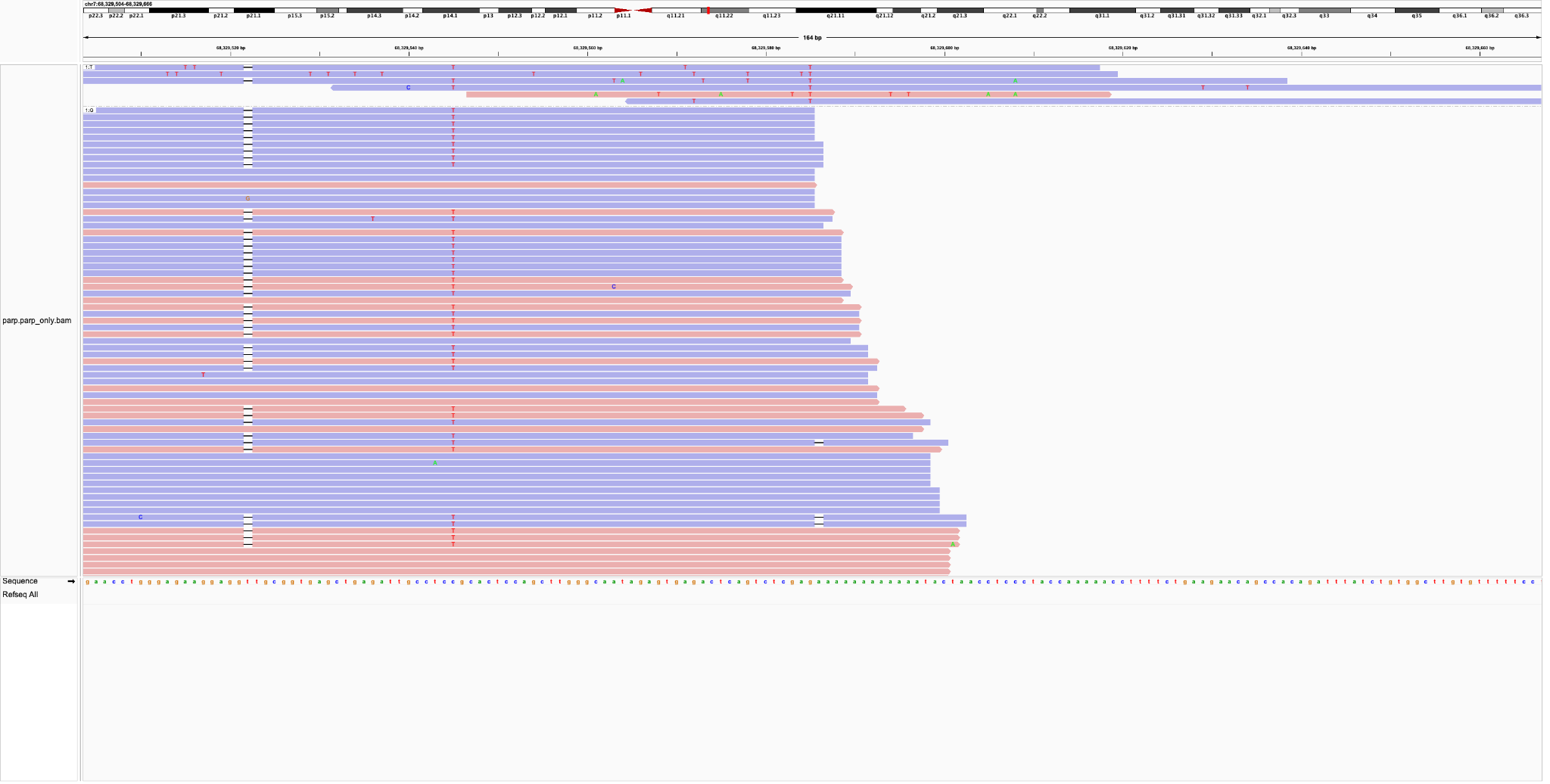

D)

E)
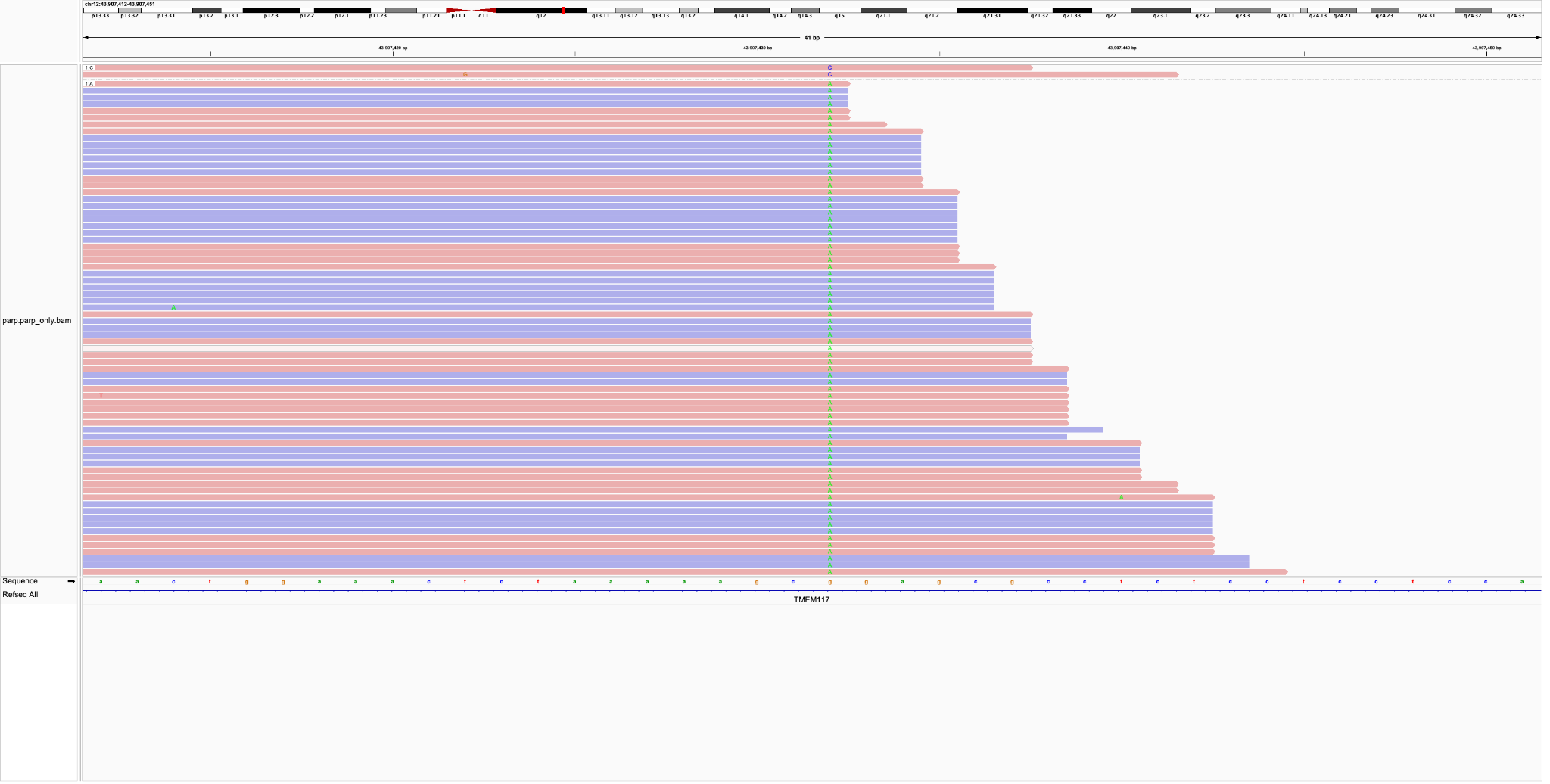

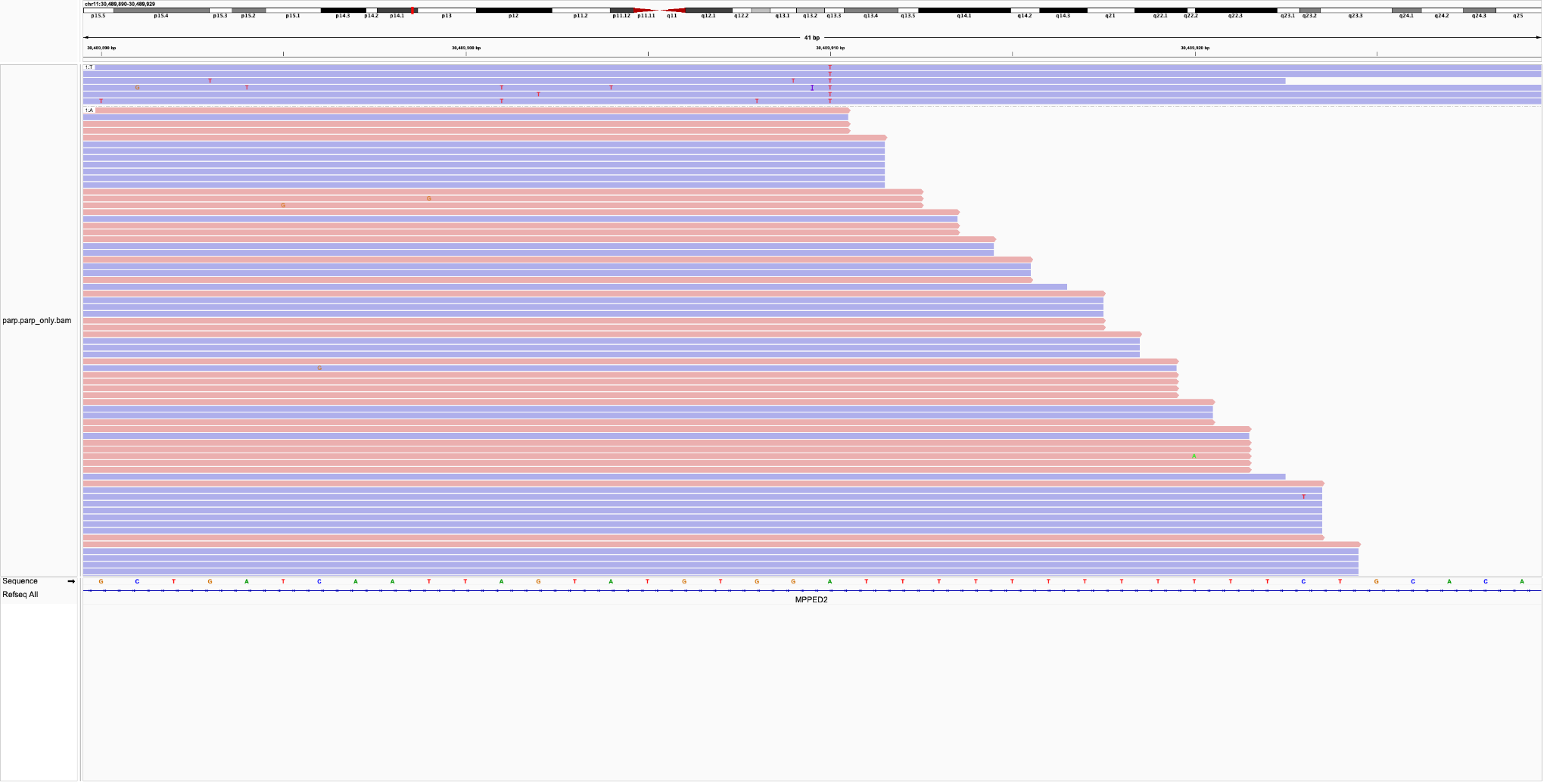

F)

***Supplementary figure S11***. Examples of false positive variants with AF > 5% that were detected in both PAPR and default pangenomes.

A) B)

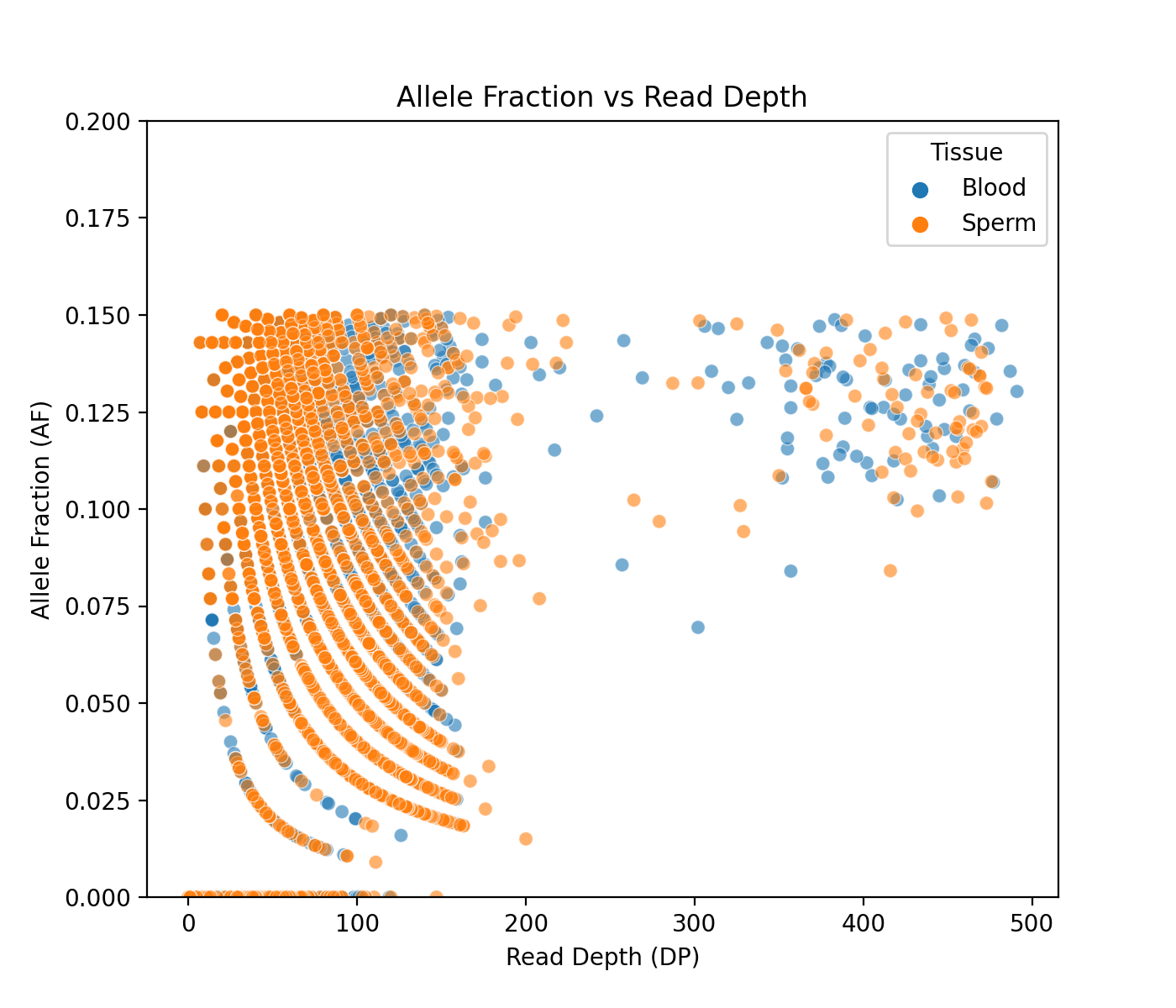

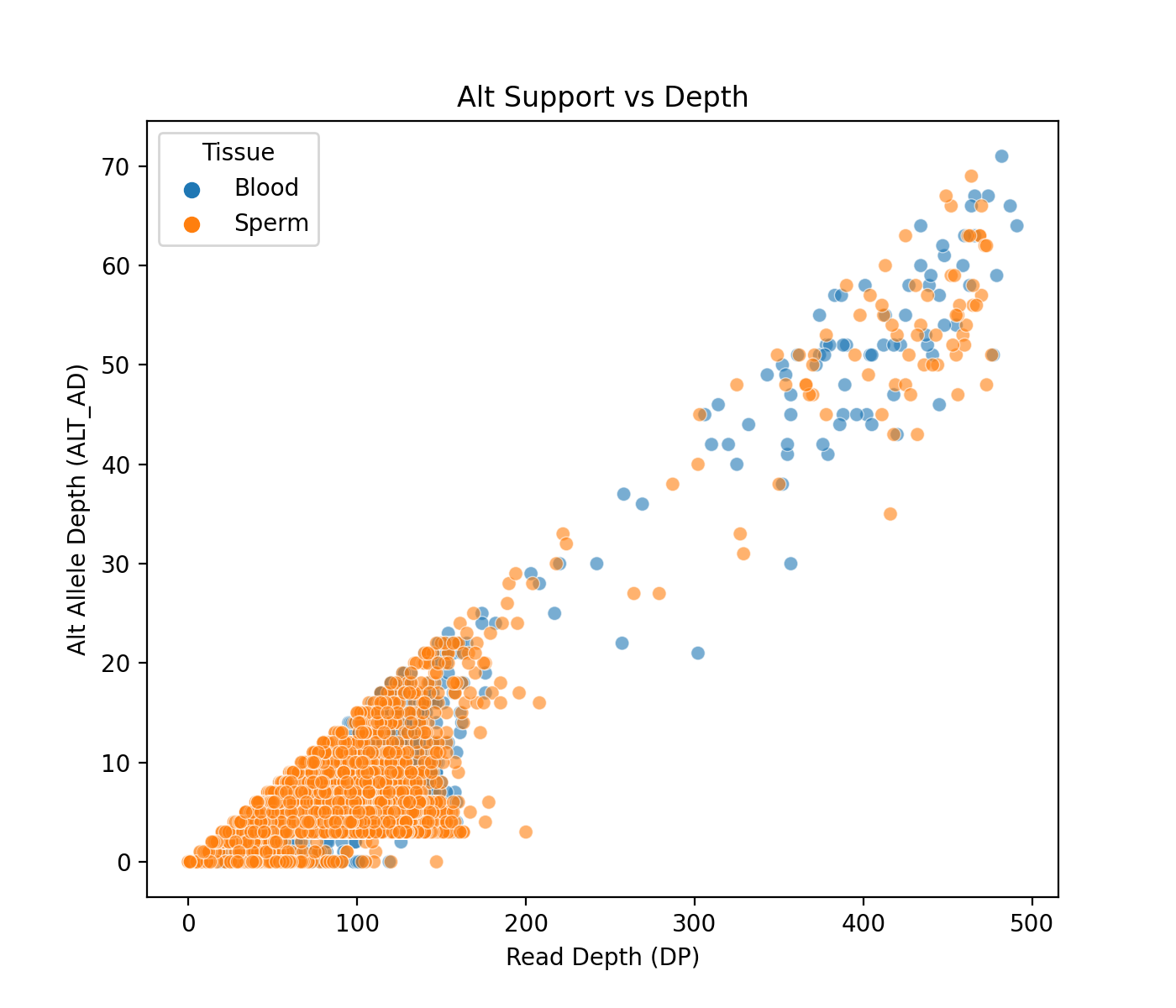

C)

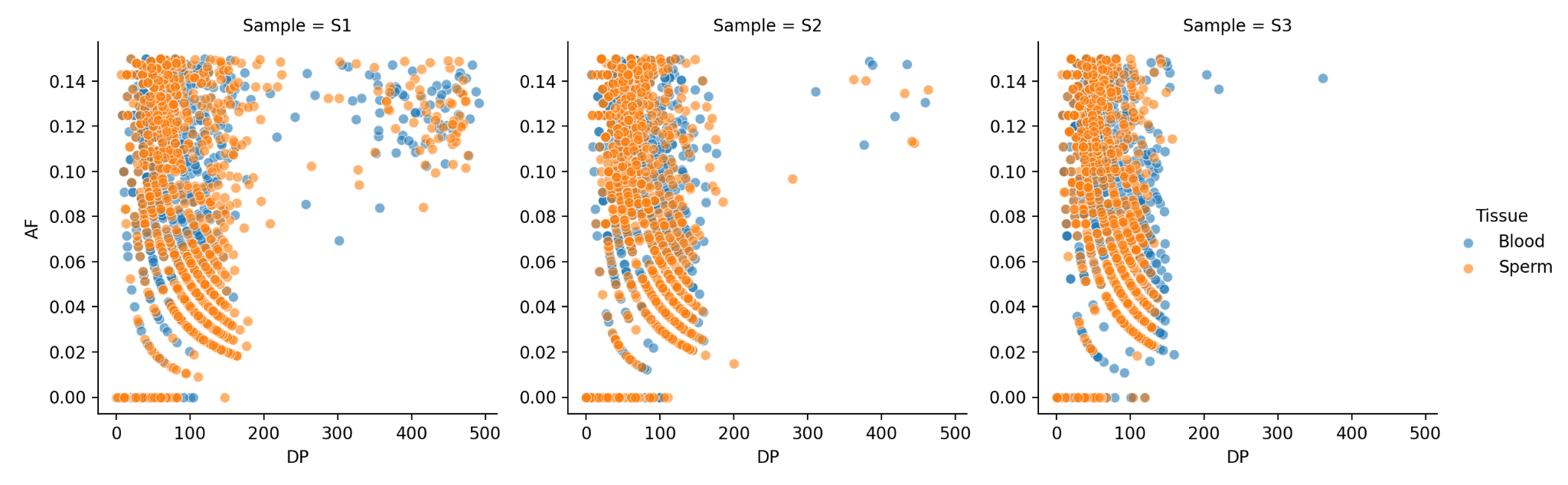

***Supplementary Figure S12*:** ***Variant allele fraction distribution across different coverage levels for three samples (~100× coverage).*** ***A)*** *AF vs read depth for all samples together* ***B)*** *Alt AD vs DP for all samples together* ***C)*** *AF vs DP for Individual samples. In sample 1, numerous variants are observed within segmental duplication (segdup) regions, as shown in the top right corner (S1). In contrast, samples 2 and 3 contain only a small number of variants in segdup regions.*

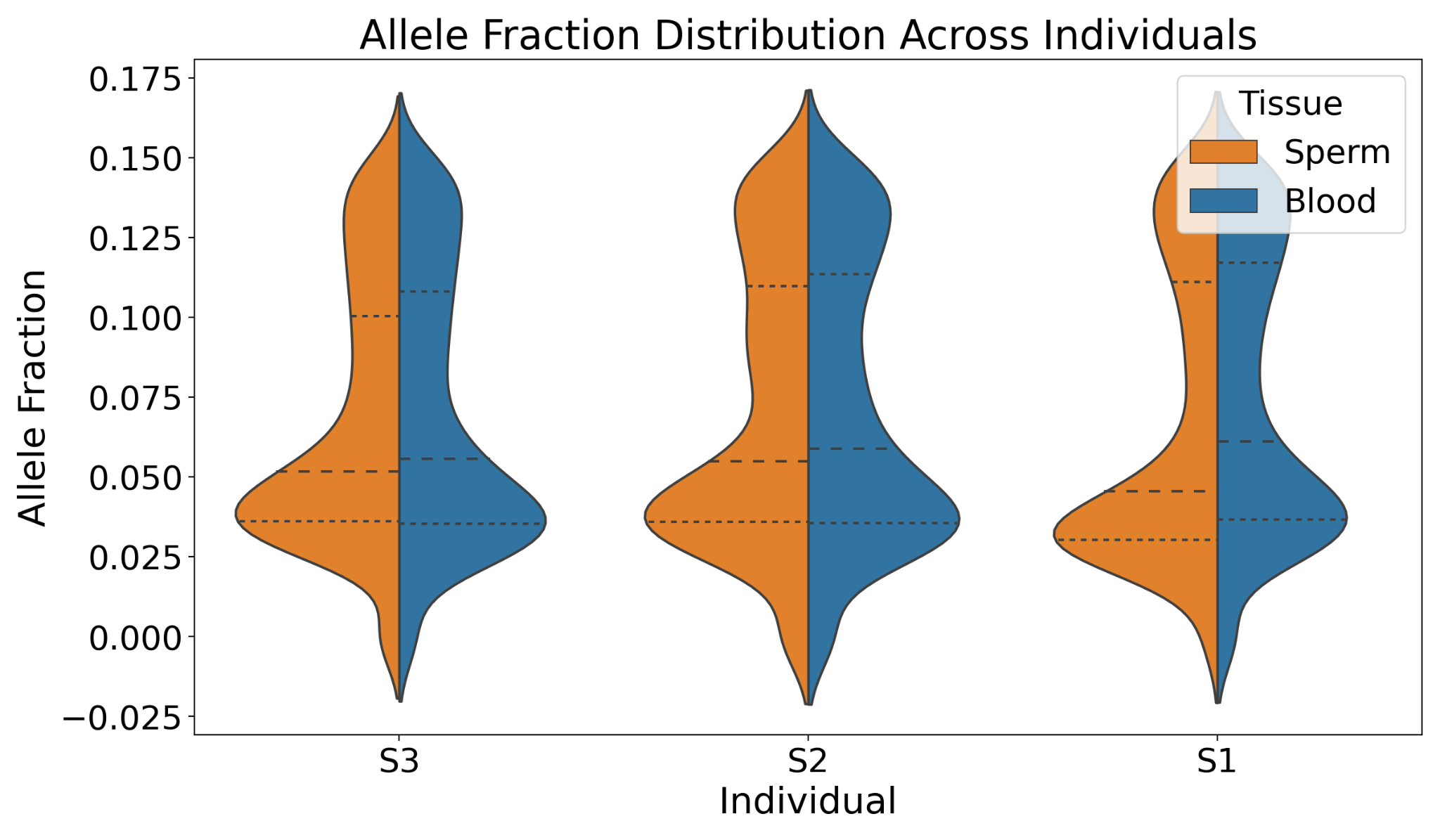

***Supplementary Figure S13:*** *The variant allele fraction distribution of low VAF variants among blood and sperm for all three samples. The sperm have more variants with VAF <=5% in the first samples, and across all three samples, the median VAF in sperm is always lower than in blood.*

A)	B)

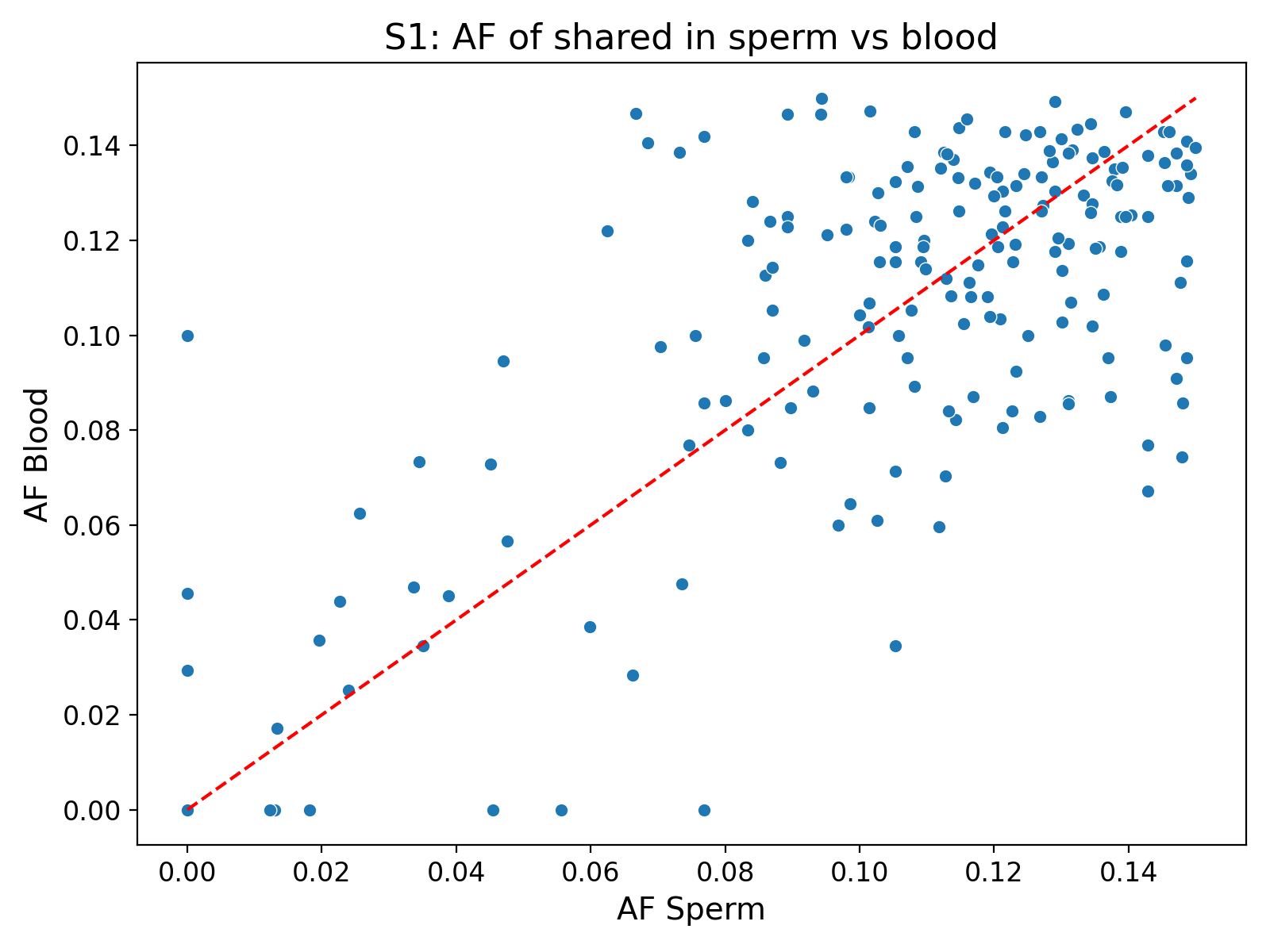

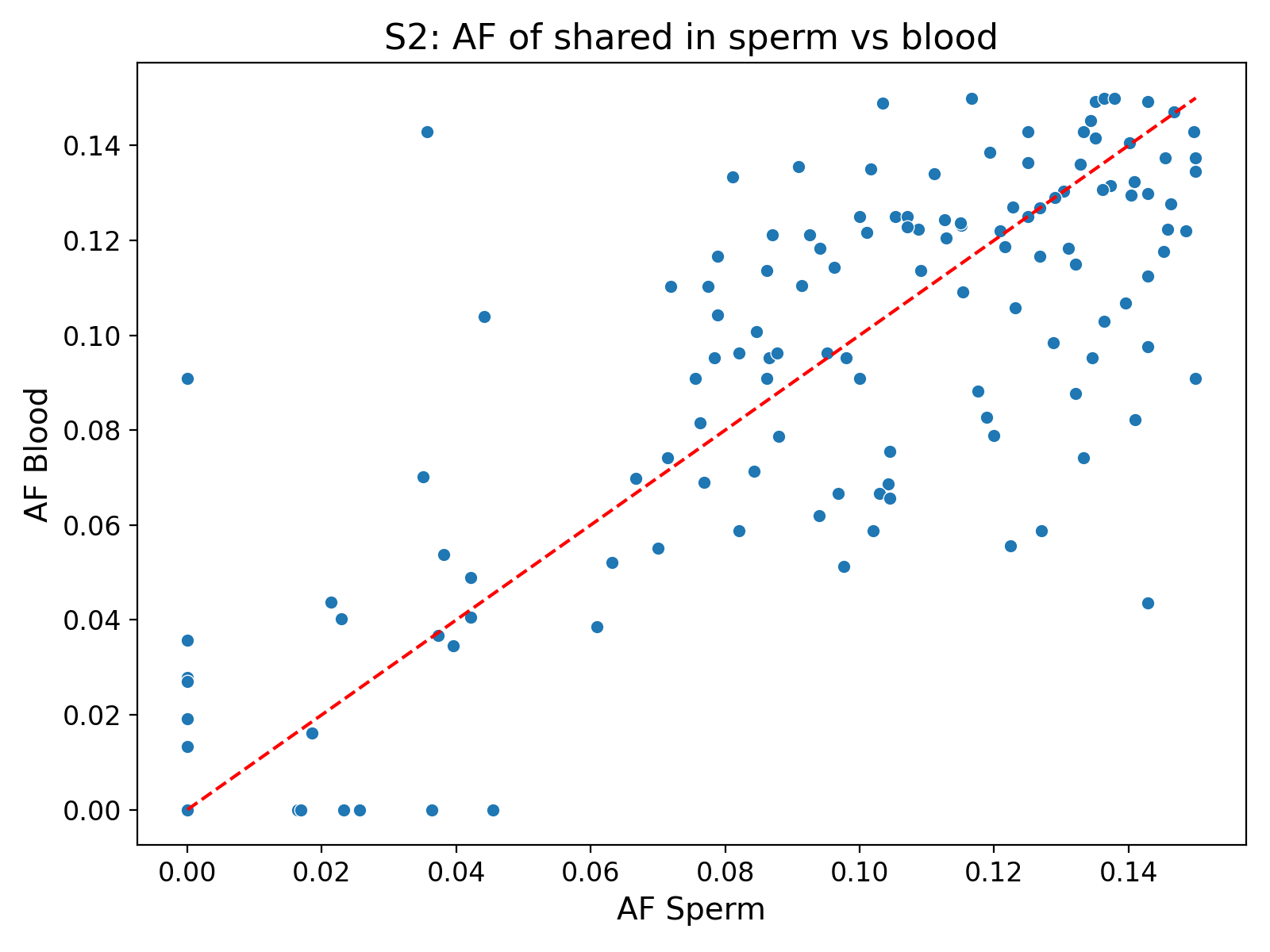

C)	D)

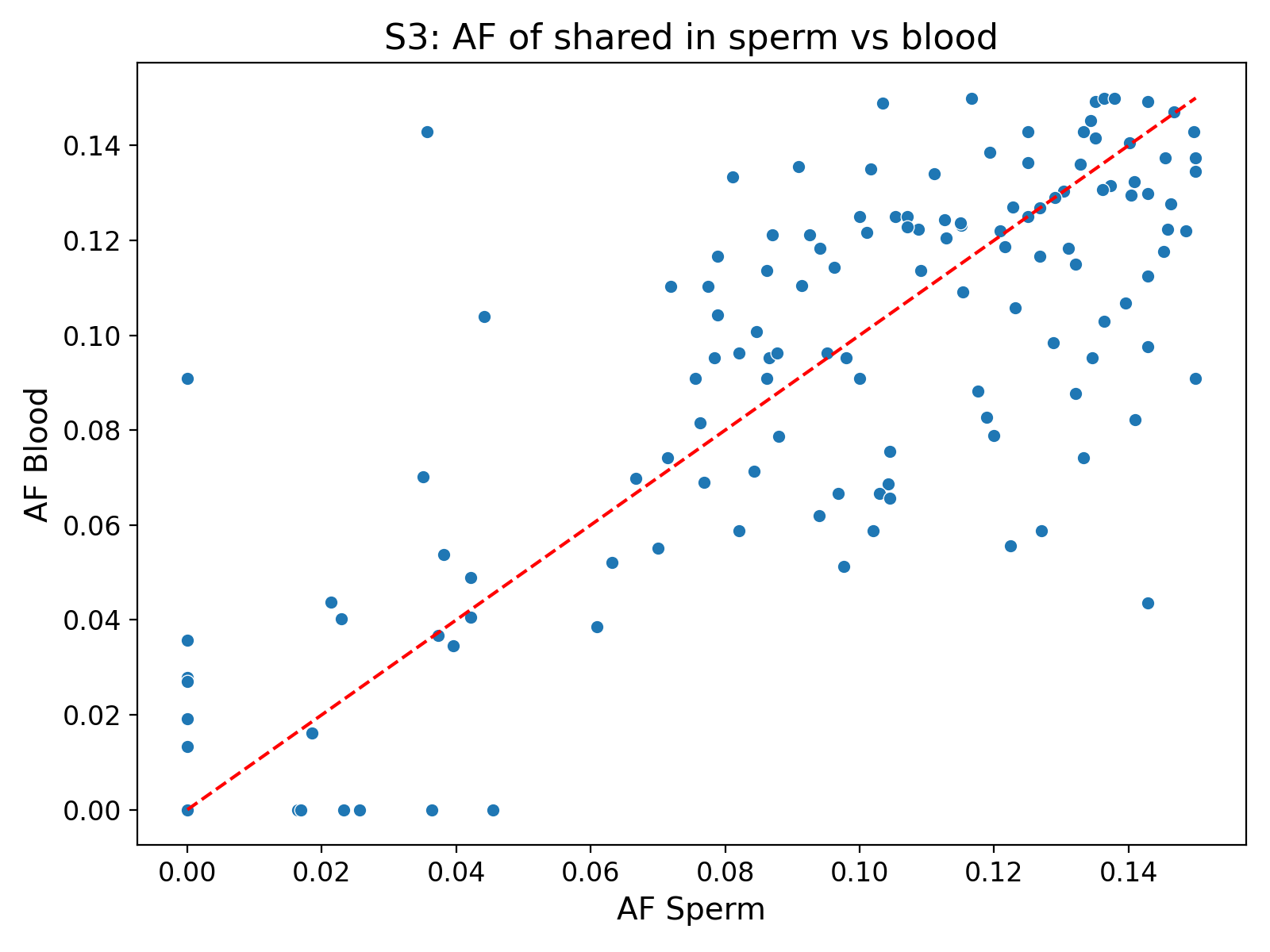

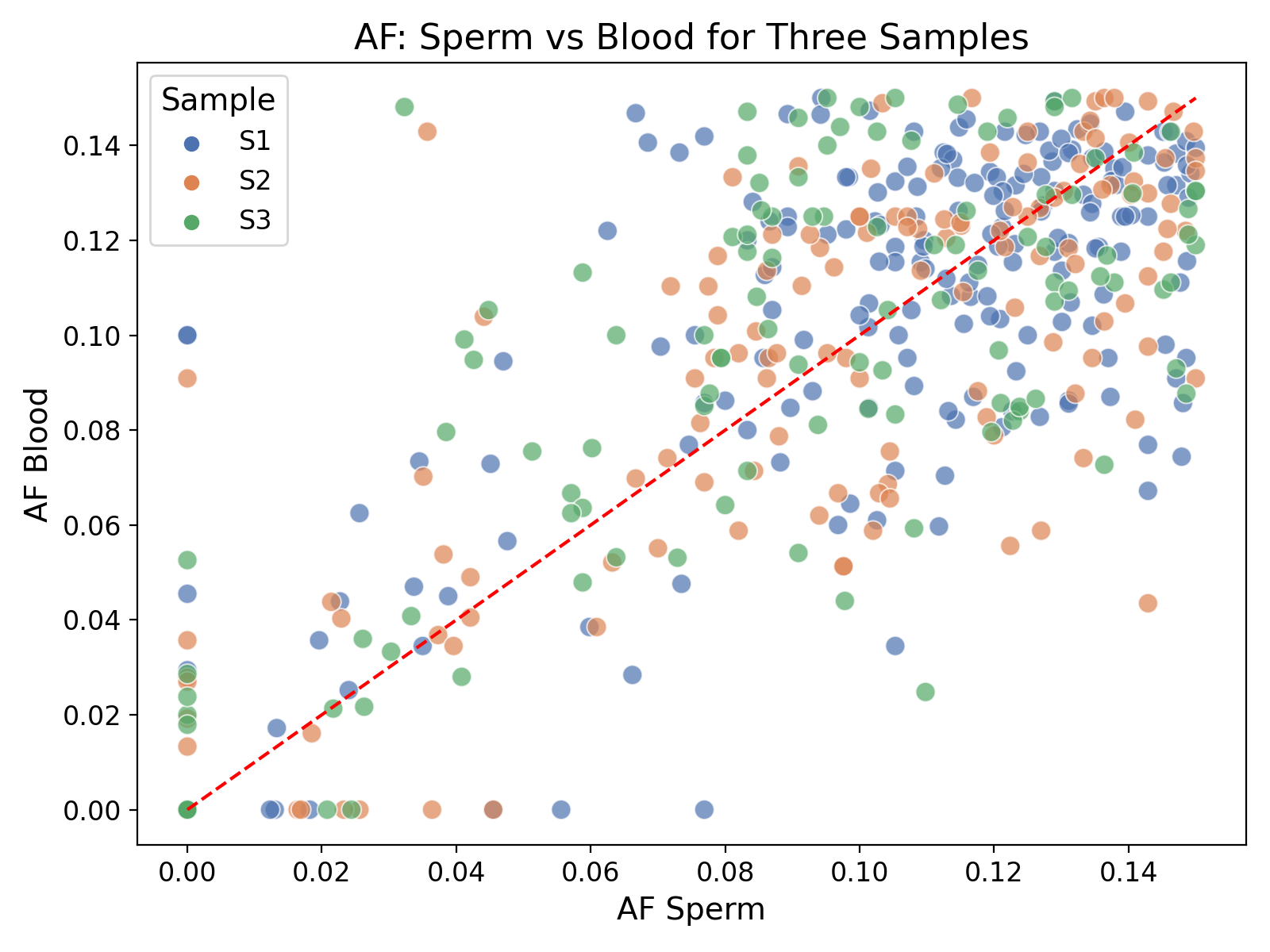

***Supplementary Figure S14*: *Shared low VAF variants:*** *AF changes at the shared positions among blood and sperm for three samples.* ***A)*** *Sample 1,* ***B)*** *Sample 2,* ***C)*** *Sample 3,* ***D)*** *all samples*

A)

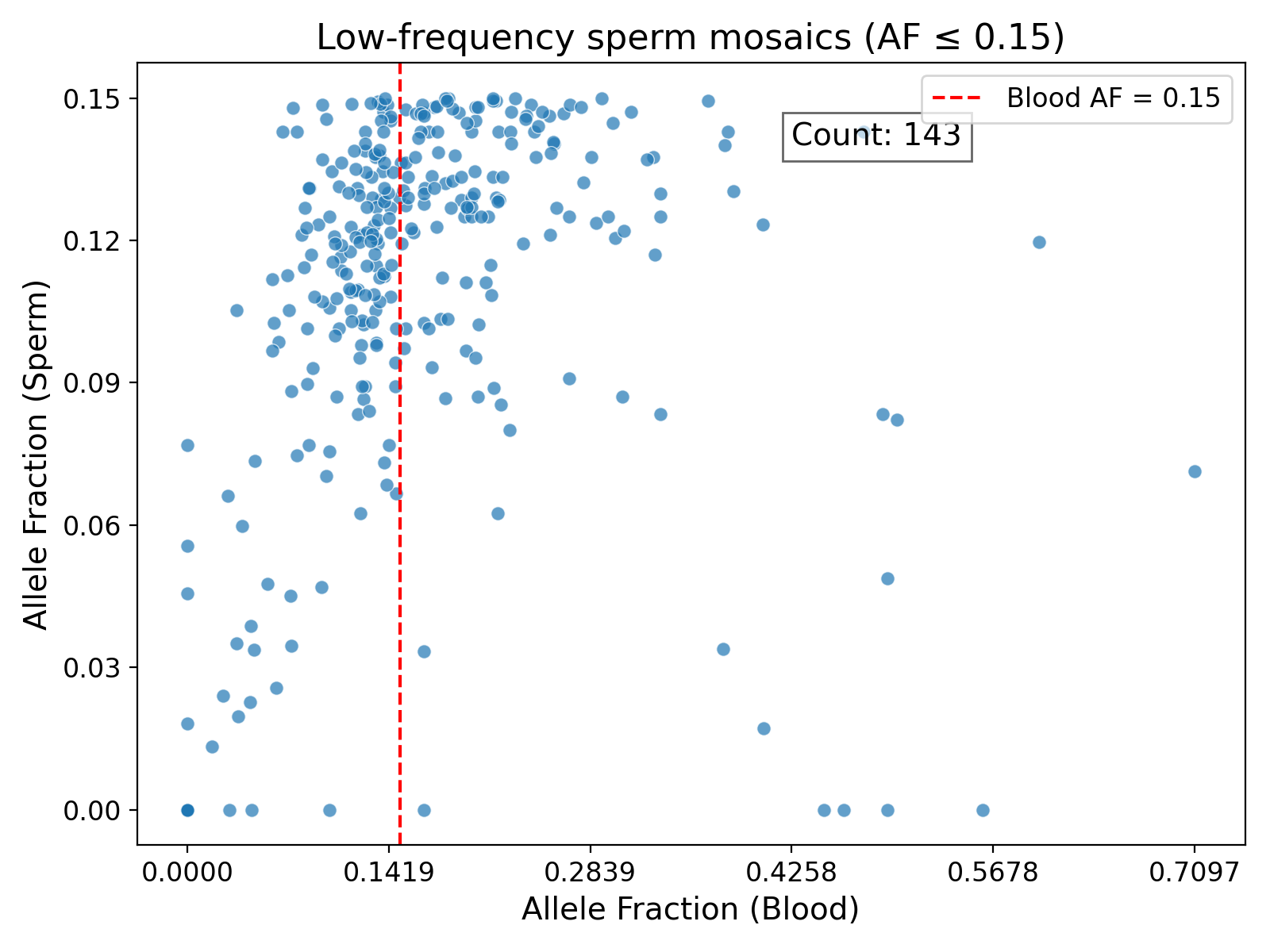

B)

C)
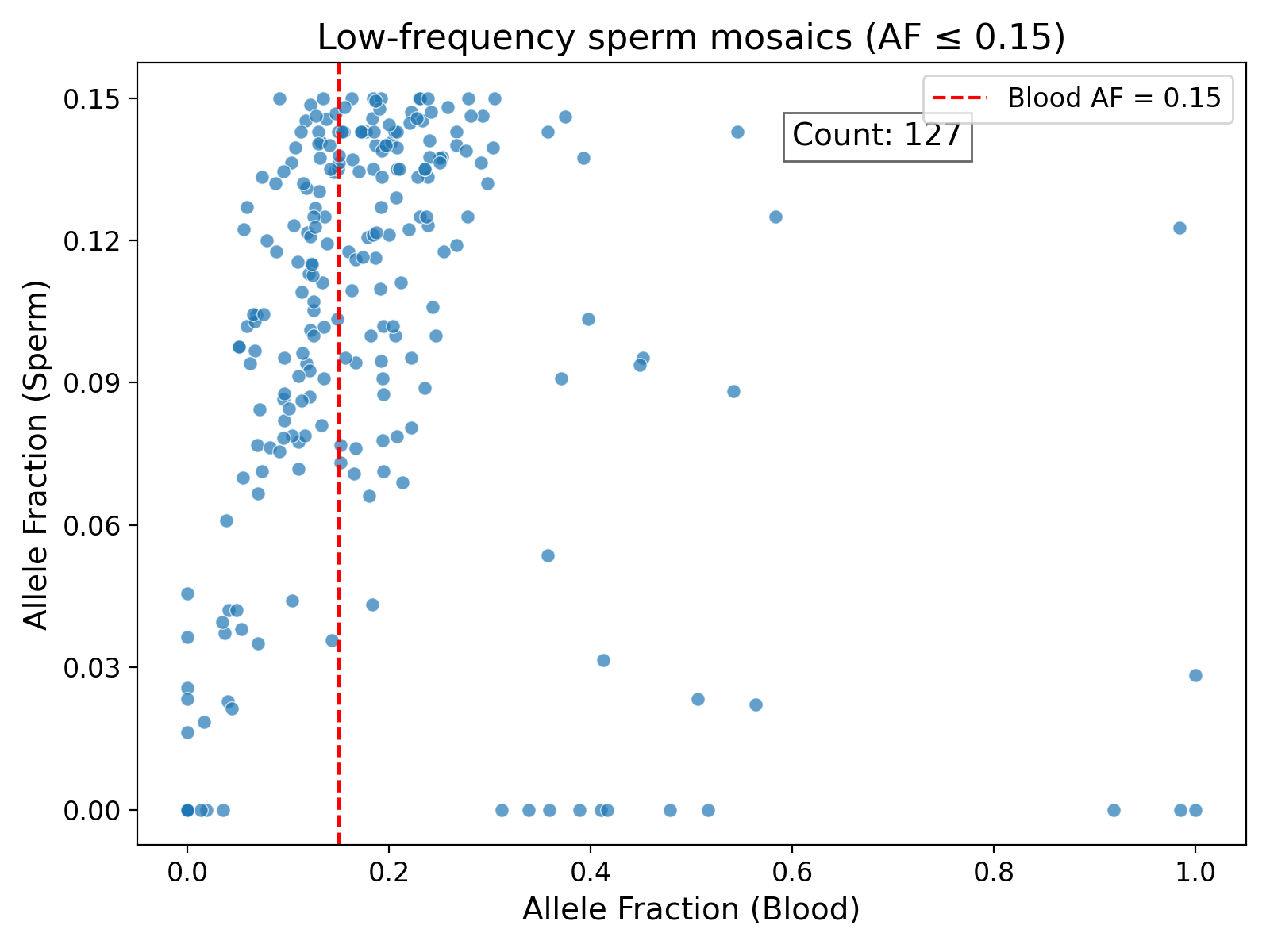

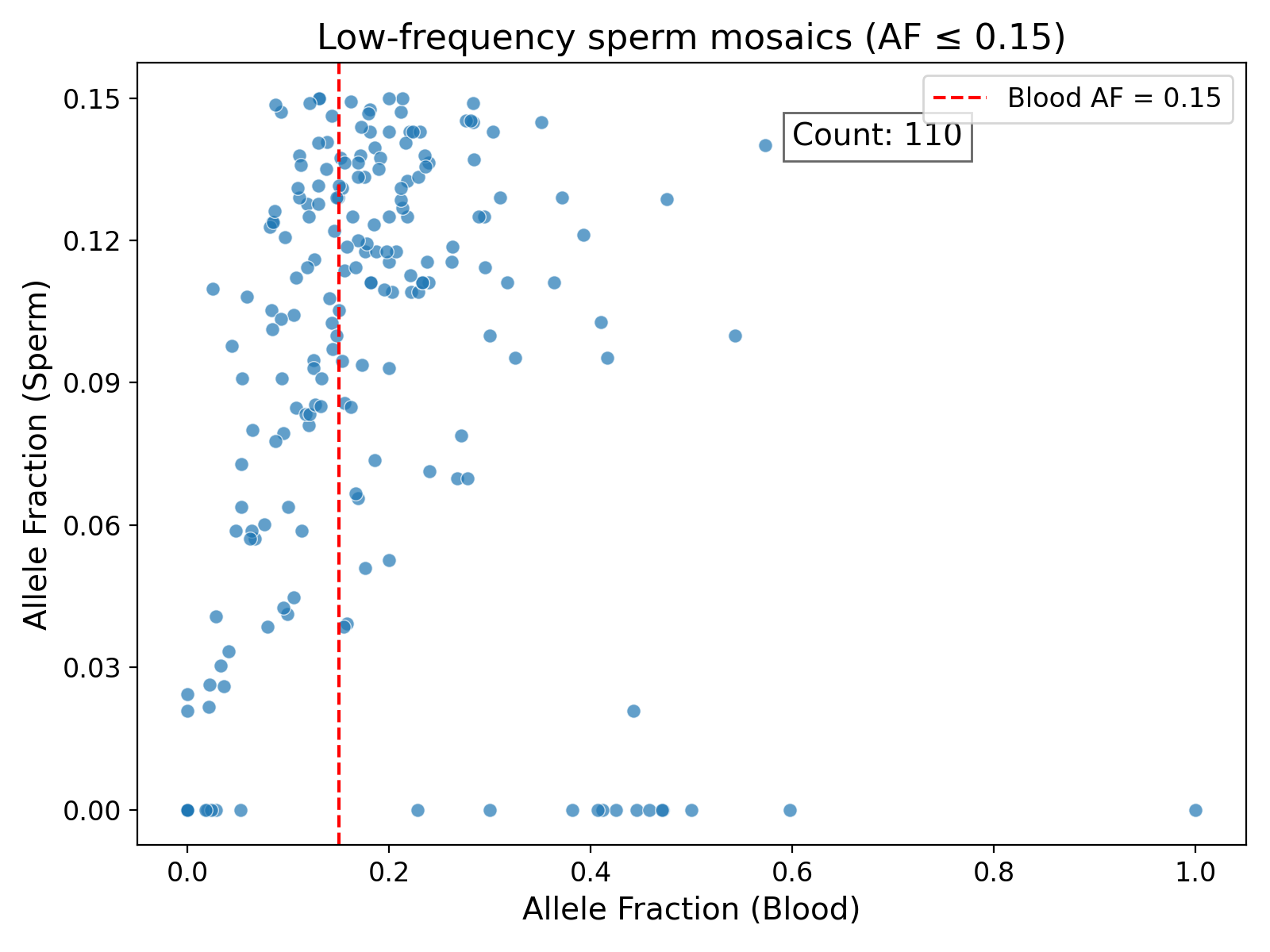

***Supplementary Figure S15*:** *Low VAF sperm variants with higher VAF in blood in* ***A)*** *Sample S1,* ***B)*** *Sample S2, and* ***C)*** *Sample S3.*

A)

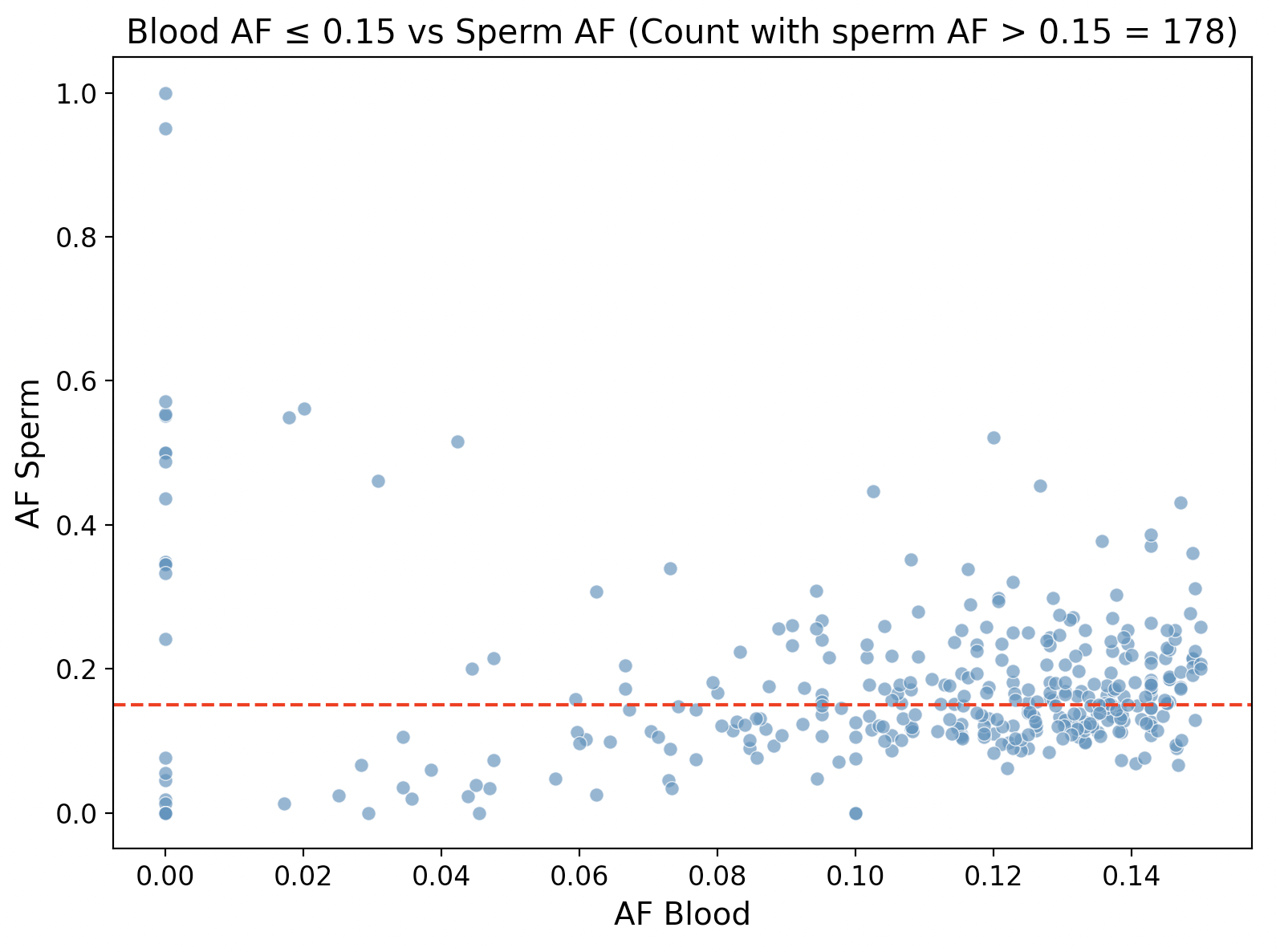

B)

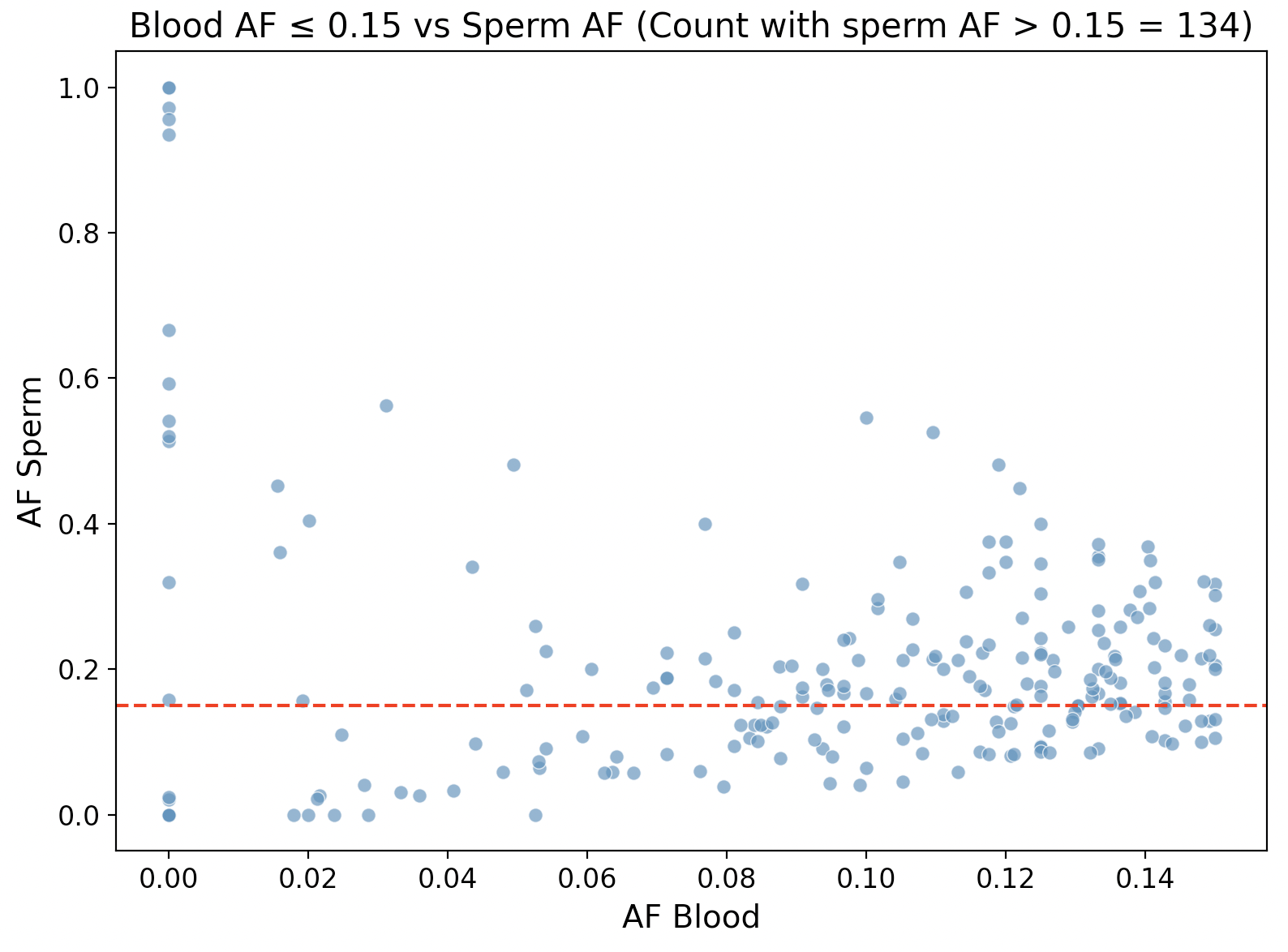

C)

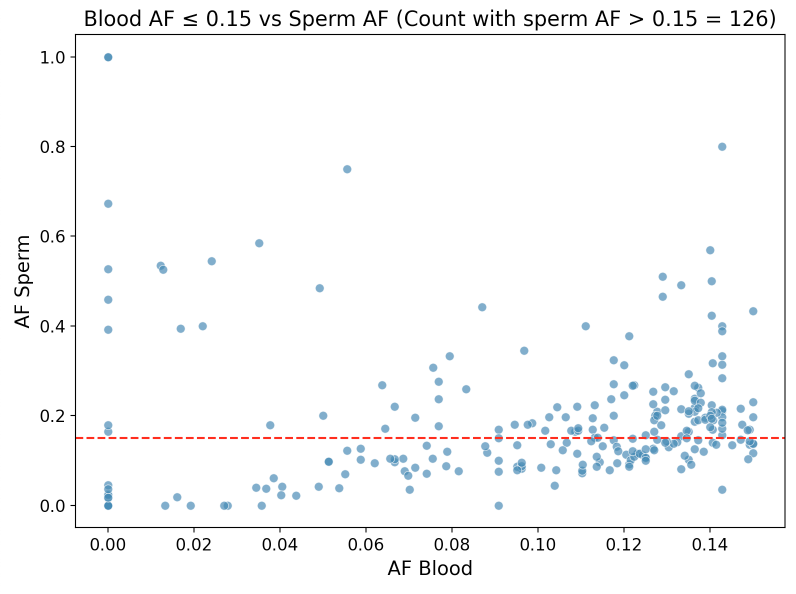

***Supplementary Figure S16*:** *Low VAF blood variants with higher VAF in sperm in* ***A)*** *Sample S1,* ***B)*** *Sample S2, and* ***C)*** *Sample S3.*

A)

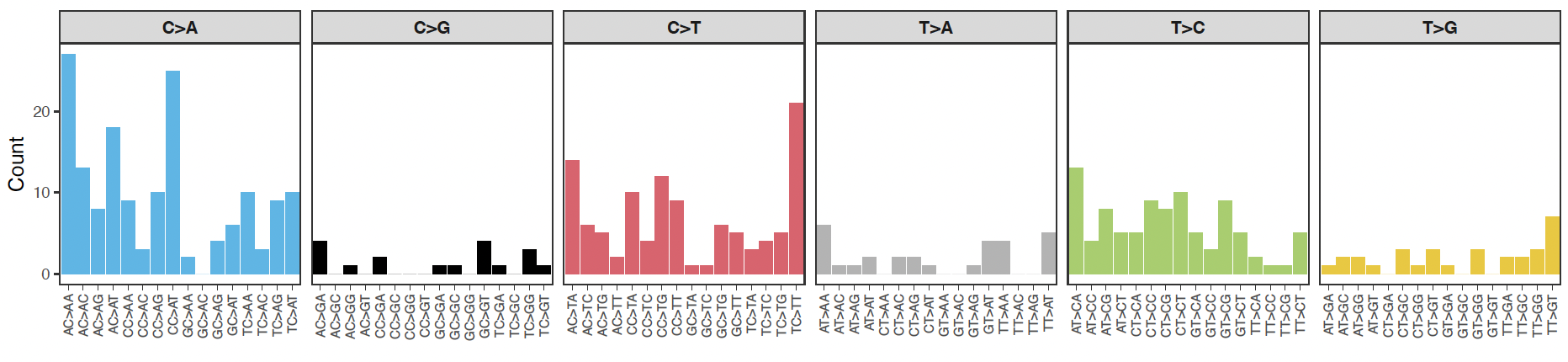

B)

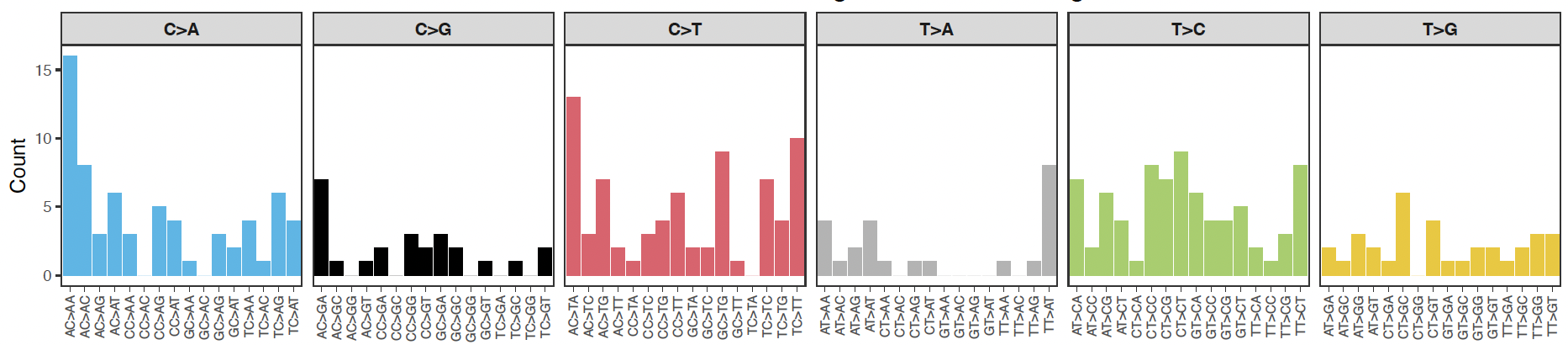

***Supplementary Figure S17:*** *The mutational signatures of A) Blood tissues and B) Sperm tissues of Sample S2.*

A)

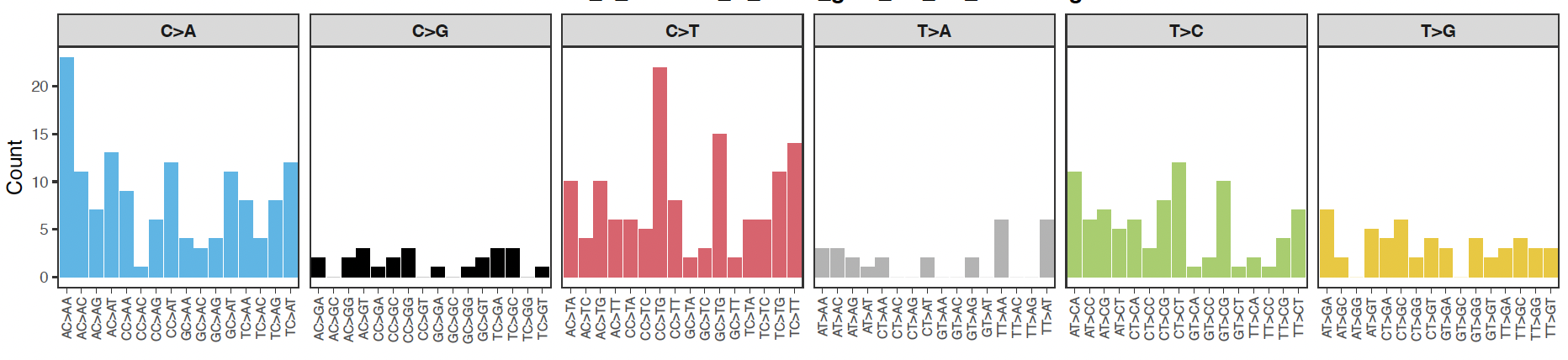

B)

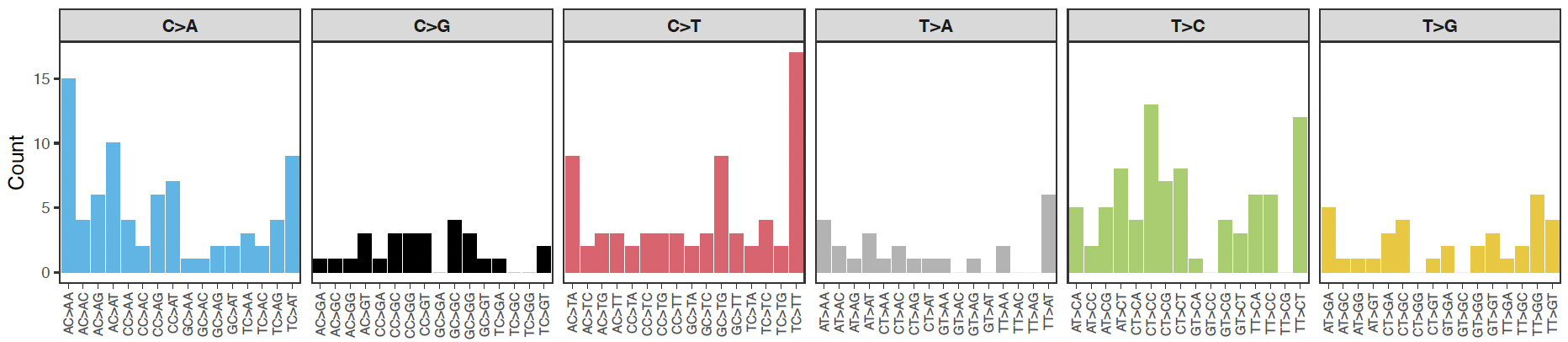

***Supplementary Figure S18:*** *The mutational signatures of A) Blood tissues and B) Sperm tissues of Sample S3.*

#### 1. Mosaic SNV benchmark generation

To assess the accuracy of mosaic variant detection down to 0.75% VAF for the HapMap mixed samples (HG005, HG001, and HG002), we generated a benchmark set by combining well-characterized NIST GIAB curated SNV benchmark^1^ (GIAB v4.2.1) sets for these samples.

The following are the GIAB v4.2.1 datasets (VCFs and high-confidence BED regions) for the three samples:

HG001: <https://ftp-trace.ncbi.nlm.nih.gov/ReferenceSamples/giab/release/NA12878_HG001/NISTv4.2.1/GRCh38/>

HG002:<https://ftp-trace.ncbi.nlm.nih.gov/ReferenceSamples/giab/release/AshkenazimTrio/HG002_NA24385_son/NISTv4.2.1/GRCh38/>

HG005:<https://ftp-trace.ncbi.nlm.nih.gov/ReferenceSamples/giab/release/ChineseTrio/HG005_NA24631_son/NISTv4.2.1/GRCh38/>

For the HapMap mix, the DNA was combined in a ratio of 60:2:1, corresponding to 95.3% HG005, 3.2% HG001, and 1.5% HG002, and this mixture mimics a combination of germline and mosaic variants, as the contributions from HG002 and HG001 result in lower VAFs, while HG005 contributes to higher VAF germline variants. The expected VAFs for homozygous variants were 3.17% for HG001 and 1.59% for HG002, whereas the expected VAFs for heterozygous variants were 1.59% and 0.79% for HG001 and HG002, respectively (**Figure 2B**). The expected VAFs for the HG005 homozygous variants were 95.238% and 47.65% for heterozygous variants . By leveraging the GIAB benchmark sets for these three individual samples, we constructed an admixed benchmark set where the variants from HG005 were considered germline variants and the variants unique to either HG001 or HG002 or both were designated as mosaic variants. To begin, the high-confidence BED regions of all three samples were intersected using bedtools(v2.30)intersect to define genomic regions shared across the samples. The GIAB v4.2.1 SNV benchmark VCF files for HG001, HG002, and HG005 were then merged with bcftools(v1.19) merge to capture all variants that are present in either one of these samples. The merged VCF file contained three sample columns with their genotype information. To reduce the complexity and simplify the benchmark set, the bi-allelic variants ( bcftools view -m2 -M2) and multi-allelic variants ( bcftools view -m3) were separated into distinct VCF files. The multi-allelic sites were excluded from the final benchmark set , thus keeping only the bi-allelic VCF file. The regions corresponding to the multi-allelic sites were also removed from the intersected BED file. The candidate mosaic variants were filtered from the bi-allelic variant file and defined as those not observed as germline in HG005. Finally, BED regions containing HG005 germline variants were filtered out from the high-confidence regions (bedtools subtract -a merged.bed -b multi_allelic.vcf > high_confidence.bed). The mosaic variants were tagged as “MOSAIC” and one more sample column (merged_hg005_01_02) was added in the final VCF file that puts GT 0/1 for mosaic variants. The distribution of benchmark mosaic variants across the HG001 and HG002 genotype combination is shown in **Supplementary Figure S2**. Altogether, this benchmark includes 3,512,786 germline and 2,208,603 mosaic variants, and it covers 80.12% of the human genome (GRCh38).

#### 2. Clonal dynamics in blood vs. sperm

To systematically assess clonal dynamics of low-VAF mosaic variants, we compiled a curated gene panel for mosaicism analysis encompassing four functional categories ^2^^,^ ^3^^,^^4,5^^,^^6^(see **Supplement Table 12**). The first category includes 14 blood-only clonal Hematopoiesis/CHIP genes, such as *DNMT3A, TET2, ASXL1,* and *JAK2*, which commonly acquire somatic mutations in hematopoietic cells due to age-related drift and clonal expansion. These variants are restricted to blood and are generally not heritable and not relevant for sperm transmission. The second category includes 31 sperm-only / Spermatogenesis-related genes, e.g., *DAZ, BOLL, SYCP1-3, TEX11-15,* *PIWIL1-4,* etc., that comprise genes essential for germ cell development and fertility. These variants in these genes could disrupt spermatogenesis or sperm function, but are less likely to cause developmental disorders. The third category contains 14 selfish spermatogonial selection/paternal age-effect genes, such as *FGFR2, FGFR3, HRAS, KRAS, NRAS, BRAF, RAF1, PTPN11,* and *SOS1*. These variants are prone to clonal expansion in aging spermatogonia. The somatic variants in these genes can be transmitted to offspring and are associated with dominant developmental disorders. The last category contains 16 high-impact de novo/developmental disorder genes that are the genes frequently found in de novo dominant disorders, such as *SCN2A, SCN1A, KCNQ2, SYNGAP1, CHD2, CHD8, ARID1A,* and *SETD2*. The mosaicism in sperm at these loci carries strong heritability and disease risk.
